# Post-discharge Oral Home Antibiotics Versus No Oral Home Antibiotics in Complicated Pediatric Appendicitis: A Systematic Review and Meta-analysis

**DOI:** 10.1101/2025.07.20.25331854

**Authors:** Javier Arredondo Montero, María Rico-Jiménez

**Affiliations:** Pediatric Surgery Department, Complejo Asistencial Universitario de León, León, Spain; Pediatric Surgery Department, Hospital Universitario Niño Jesús, Madrid, Spain

**Keywords:** complicated acute appendicitis, pediatric, oral antibiotics, discharge, surgical site infection, readmission, stewardship

## Abstract

**Background:** The use of oral home antibiotics (OHA) after discharge in children undergoing surgery for complicated acute appendicitis (CAA) remains controversial. This systematic review and meta-analysis aimed to evaluate whether OHA reduces the risk of infectious complications or readmissions compared to patients discharged without antibiotics (NHA).

**Methods:** This systematic review was prospectively registered in PROSPERO (CRD420251049919). We searched PubMed, Web of Science, Scopus, Ovid, and Cochrane CENTRAL from inception to March 2025. Two independent reviewers screened the studies, extracted the data, and assessed the methodological quality using the ROBINS-I tool. Eight random-effects meta-analyses and four leave-one-out meta-analyses were conducted for intra-abdominal abscesses (IAA), surgical site infections (SSI), organ/space infections (OSI), and hospital readmissions (RA). Two exploratory random-effects meta-regression models were performed for RA. Certainty of evidence for all outcomes was formally graded using GRADE.

**Results:** Fourteen studies comprising 26,174 pediatric patients with CAA were included. Meta-analyses showed no significant differences between intervention (IG) and comparator (CG) groups for IAA (RR 1.23; 95% CI: 0.62-2.45), OSI (RR 1.19; 95% CI: 0.73–1.93), or RA (RR 1.07; 95% CI: 0.78-1.44). In exposure-restricted analyses, NHA patients had a modestly lower risk of readmission (RR 0.78; 95% CI 0.61–1.01; p = 0.05), suggesting a potential harmful signal for OHA. The risk of SSI was significantly higher among patients in the CG (RR 0.77; 95% CI, 0.61–0.96; p = 0.02). However, this apparent association was not robust and was lost in sensitivity analyses restricted to studies with crude patient-level exposure data, where the effect reversed direction (RR > 1), consistent with protocol-based confounding. Across all outcomes, certainty of evidence was rated very low, primarily driven by potential confounding by indication and non-randomized designs.

**Conclusions:** OHA after discharge does not appear to reduce the risk of postoperative complications in children treated surgically for CAA. Given the lack of consistent benefit and potential for unnecessary harm, routine use of post-discharge OHA is not supported. The exposure-restricted analysis also raises a plausible signal of harm in terms of RA. Because the certainty of evidence is very low, further high-quality prospective research is needed to clarify the true effect of OHA in this context.

**Funding:** None.

**Registration:** PROSPERO (CRD420251049919).

## Introduction

Acute appendicitis (AA) is the most common surgical emergency in the pediatric population, with an estimated annual incidence of 9.68 per 10,000 children [1]. While most cases present as non-complicated appendicitis (NCAA), approximately 15–25% present as complicated acute appendicitis (CAA) [2,3]. Patients with CAA are more susceptible to postoperative complications and typically require prolonged antibiotic therapy and extended hospitalization, contributing to a substantial burden on healthcare systems and resource utilization [4]. While CAA generally includes gangrenous or perforated forms, it is crucial to outline that the specific operational definitions vary considerably across the literature. This heterogeneity constitutes a key challenge in synthesizing the data, and a detailed examination of the criteria used by each included study is a component of this review.

Despite consensus on the need for intravenous antibiotic therapy during hospitalization for CAA, and a growing trend toward institutional protocols promoting shorter intravenous courses or combined intravenous–oral regimens—with outcomes shown to be non-inferior to traditional approaches [5,6]—substantial variation persists in post-discharge management, particularly regarding the use of oral antibiotics. Current guidelines provide limited or no specific recommendations on antibiotic continuation after discharge, leading to heterogeneous institutional practices and variability among surgeons. Several studies have reported marked inter-professional variability in this aspect of care, underscoring both the lack of standardization and the ongoing uncertainty regarding its clinical benefit [7].

The rationale for prescribing oral antibiotics after hospital discharge in cases of CAA is primarily based on the assumption that continued antimicrobial coverage may reduce the risk of delayed infectious complications, such as surgical site infections (SSI), intra-abdominal abscesses (IAA), organ space infections (OSI), or unplanned readmissions (RA). Concerns over early hospital discharge and residual contamination in perforated or gangrenous appendicitis have traditionally justified this strategy. Oral home antibiotics (OHA) are often perceived as a low-cost, accessible intervention that may provide a safety buffer during the vulnerable post-discharge period.

However, prolonged antibiotic exposure is not without risks. The unnecessary use of antimicrobials contributes to adverse drug reactions, disruption of the gut microbiota, an increased risk of *Clostridioides difficile* infection, and a broader public health threat from antimicrobial resistance [8,9]. Given their developing immune and microbial systems, these concerns are particularly relevant in pediatric populations. Notably, emerging evidence suggests that post-discharge oral antibiotic therapy may offer limited or no benefit in reducing complications—and may even be associated with overtreatment and an increased risk of adverse events.

Given the widespread but inconsistent use of post-discharge OHA in children with surgically treated CAA—and the growing body of evidence questioning their effectiveness—there is a pressing need to synthesize the available data to inform clinical decision-making. The absence of clear guideline recommendations and the potential for both overuse and harm further underscore this gap. Therefore, the objective of this systematic review is to evaluate the impact of oral antibiotic therapy after hospital discharge on outcomes in pediatric patients with surgically treated CAA, with a specific focus on infectious complications (including SSI, OSI, and IAA) and readmissions.

## Methods

### Literature Search and Selection

We followed the Preferred Reporting Items for Systematic Reviews and Meta-Analyses (PRISMA) guidelines [10] and the Cochrane Handbook for Systematic Reviews of Interventions (version 6.5) [11]. The PRISMA checklist is provided in Supplementary File 1. This review was prospectively registered in the International Prospective Register of Systematic Reviews (PROSPERO ID: CRD420251049919).

Eligible studies were identified by searching the primary existing medical bibliography databases (PubMed, Web of Science, Scopus, Ovid, and Cochrane Central). Supplementary File 2 shows the detailed search strategy for each bibliographic database. The search was last executed on 11.05.2025.

JAM and MRJ selected articles using the COVIDENCE ® tool. The search results were imported into the platform, and both authors independently screened the articles. Disagreements were resolved by consensus. Supplementary File 3 shows the inclusion and exclusion criteria.

### Quality Assessment

The Cochrane Risk of Bias 2 (RoB 2) tool was selected to evaluate the methodological quality and risk of bias of randomized controlled trials (RCTs) [12]. In contrast, the Risk of Bias in Non-randomized Studies - of Interventions (ROBINS-I) tool was chosen to assess the methodological quality and risk of bias of non-randomized studies of interventions (NRSIs) [13]. No RCTs were ultimately identified during study selection; therefore, RoB2 was retained only as a pre-specified protocol element and was not applied in practice.

### Data Extraction and Synthesis

Two reviewers (JAM, MRJ) independently extracted data from the included studies using a standardized and piloted form.

Extracted variables included: first author, year of publication, country of study, study design, sample size, age, sex distribution, Intervention and comparator groups definition, discharge criteria, total leukocyte count (TLC) admission values, TLC predischarge values, antibiotics employed (type, duration, and route), events by group including SSI, IAA, OSI, RA, reoperations (RO), and visits to the Emergency Department (EDV), and p-values for between-group comparisons. Given the known heterogeneity in defining CAA, a pre-specified variable for extraction was the explicit criteria used by each study. Data on the definition of CAA were extracted and categorized, including criteria based on intraoperative findings such as perforated acute appendicitis (PAA) identified as a “visible hole in the appendix”, the presence of a free fecalith in the abdominal cavity, the presence of an intra-abdominal abscess, diffuse fibrinopurulent exudate, contamination extending beyond the right lower quadrant or involving more than two quadrants, diffuse peritonitis, intra-abdominal stool, or endoluminal content leaking during surgery. We also extracted whether the definition included gangrenous appendicitis (GA) or was based on histopathological confirmation. Finally, we noted whether studies applied mixed criteria or did not specify the criteria used.

Disagreements during extraction were resolved by discussion or consultation with a third reviewer. Study authors were contacted for clarification when critical data were missing or unclear. In studies where raw data were available disaggregated by treatment group — OHA versus no home antibiotics (NHA) — these values were directly used; when not available, they were inferentially estimated, and if this was not possible, events were reported according to treatment protocol groups (e.g., “old protocol” versus “new protocol”), explicitly stating the lack of specific data for OHA and NHA. Unit standardization was not required.

### Meta-analysis

Four primary random-effects meta-analyses including all eligible studies, and four sensitivity analyses restricted to studies reporting crude patient-level exposure data were conducted using the restricted maximum likelihood (REML) estimator for both the pooled effect size and the between-study variance (τ²), following the Cochrane Handbook recommendations (version 6.5) [11, 14–16]. For each outcome, the choice of confidence interval (CI) around the pooled effect was based on the number of included studies and the τ². When τ² was close to zero or only two studies were available, conventional Wald-type intervals were deemed acceptable. When more than two studies were included and τ² was greater than zero, CIs were computed using both the Hartung–Knapp–Sidik–Jonkman (HKSJ) [15] and the truncated modified Knapp–Hartung (mKH) methods [16]. The approach providing the most robust and interpretable inference was selected. For all primary random-effects models, 95% prediction intervals (PIs) were also calculated to quantify the expected range of true effects in future comparable settings. Because some studies reported outcomes based on institutional protocols rather than individual antibiotic exposure, we prospectively planned a sensitivity analysis stratifying studies by comparator structure (protocol-based vs. patient-level crude exposure OHA vs. NHA). This distinction was treated as a methodological moderator and explored in a pre-specified sensitivity meta-analysis. Exploratory leave-one-out sensitivity analyses were conducted for each one of the primary meta-analyses to assess the influence of individual studies on overall estimates. Where sufficient studies (≥10) were available, pre-specified univariable random-effects meta-regression analyses were performed to explore potential sources of heterogeneity. For studies with zero events in either treatment arm, a modified Haldane–Anscombe continuity correction was applied.

### Publication Bias and Small Study Effects

In meta-analyses including at least ten studies, publication bias and small-study effects were explored through visual inspection of contour-enhanced funnel plots, which included significance contours at conventional thresholds (p < 0.01, p < 0.05, p < 0.10, and p > 0.10), as well as through Egger’s and Begg’s statistical tests [17,18]. To complement these assessments, Galbraith (radial) plots were generated to illustrate the contribution of each study to overall heterogeneity, and L’Abbé plots were used to visually examine the distribution and directionality of individual study effects across treatment groups, rather than heterogeneity *per se*. When funnel plot asymmetry was observed, the trim-and-fill method was applied to estimate the potential impact on the pooled effect size [19].

### GRADE Assessment

The certainty of evidence for each primary outcome was evaluated using the GRADE framework. Because all included studies were non-randomized studies of interventions (NRSIs), certainty ratings started at **low**. Five domains were examined for potential downgrading: (1) risk of bias (ROBINS-I), (2) inconsistency (statistical heterogeneity: *I*^2^, τ^2^ plus visual inspection of forest plots), (3) indirectness (differences in PICO alignment), (4) imprecision (95% CI width and whether it crossed clinically relevant decision thresholds), and (5) publication bias (funnel plot asymmetry and statistical testing when n ≥ 10 studies). Potential upgrading domains (large effect size, dose–response gradient, and residual confounding favoring the null) were considered but were not applicable. Final certainty ratings were categorized as high, moderate, low, or very low, with justifications presented in the Summary of Findings table.

## Results

### Summary of the Included Studies

The search yielded 331 articles (Scopus, n = 93; PubMed, n = 57; Web of Science, n = 106; Ovid, n = 12; Cochrane CENTRAL, n = 63). After removing 150 duplicates, 181 records remained. Of these, 167 were excluded based on the predefined inclusion and exclusion criteria. Ultimately, this systematic review included 14 studies encompassing data from 26,174 patients (15,488 males, 10,686 females) [20–33]. Minimal discrepancies (<0.5%) were identified between the numbers of patients included per group (IC and CG) and per gender (male/female) reported in the review, attributable to the lack of explicit reporting in some studies. Concerning study design, no randomized controlled trials were identified; all included studies were observational. The flowchart of the search and selection process is shown in Figure 1.

**Figure 1.**
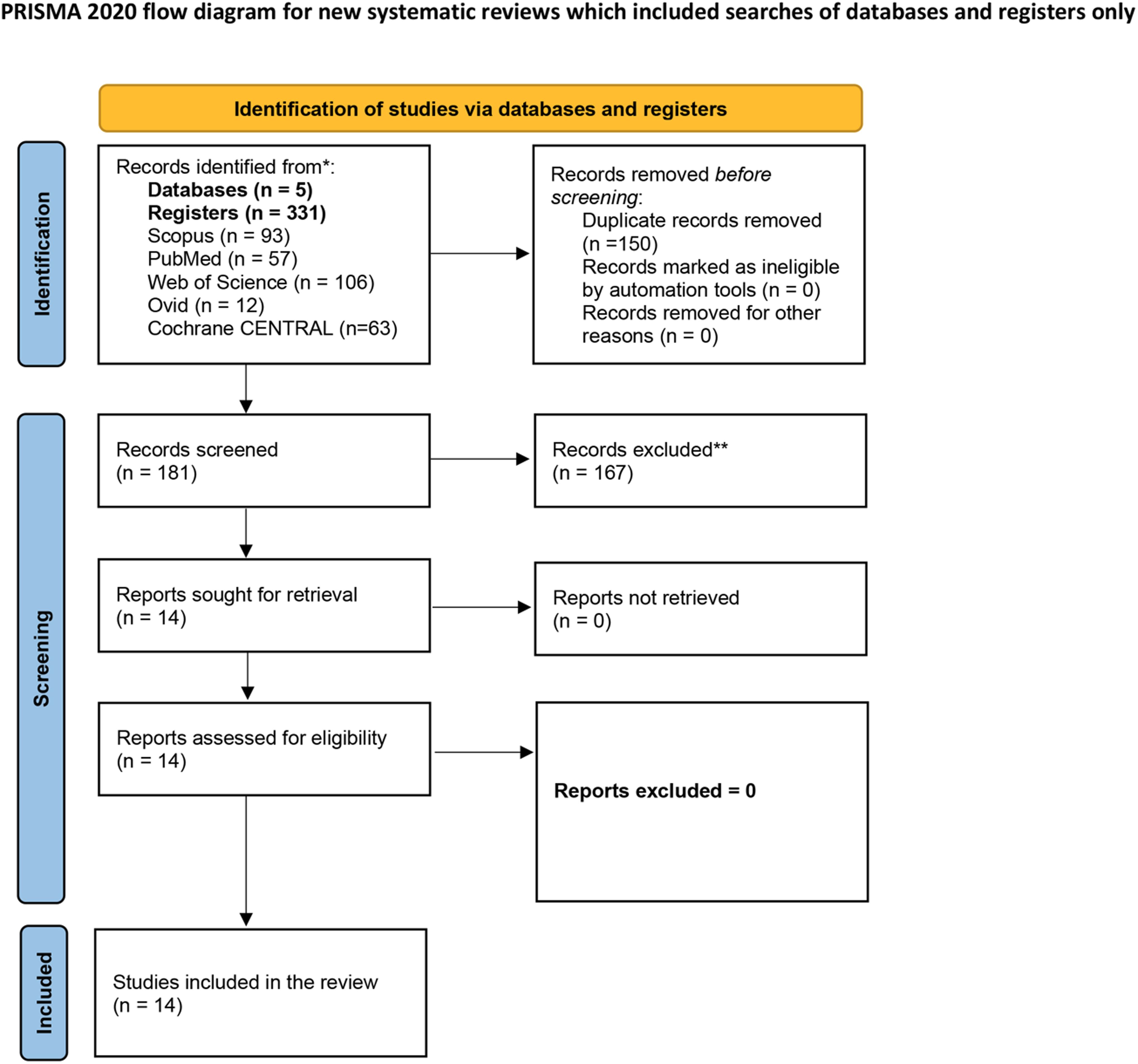
Flowchart illustrating the search strategy and study selection process.

### Sociodemographic and Clinical Characteristics

All studies were conducted between 2015 and 2025 in pediatric populations [20–33]. Thirteen were from the United States [20–29,31–33], and one was from Japan [30]. One prospective cohort study was identified [20], along with eight retrospective cohort studies [22–27, 29, 31], and five studies based on national databases, including the NSQIP Pediatric Database [21, 28, 32, 33] and the Diagnosis Procedure Combination Database [30].

All included studies focused on patients with complicated acute appendicitis (CAA), although moderate variability in the definitions used was observed. Several authors defined CAA as PAA identified intraoperatively as a “visible hole in the appendix” [20–26,28,29,31–33], or as the presence of free fecalith in the abdominal cavity [20,21,23,24,28,31–33]. Others considered CAA to include the presence of intra-abdominal abscess [21,23,28,31–33], diffuse fibrinopurulent exudate within the peritoneal cavity [21,33], fibrinopurulent contamination extending beyond the right lower quadrant and pelvis [28], or involving more than two quadrants, including the pelvis [32]. Additional criteria used were the presence of diffuse peritonitis [23], intra-abdominal stool [24], and endoluminal content leaking outside the bowel during surgery [27]. Some authors also included gangrenous appendicitis (GA) under the CAA definition [22] or based the diagnosis on histopathological confirmation of PAA [29]. One study did not specify the criteria used to define CAA [30]. It is worth noting that many studies applied mixed criteria for defining CAA, and only a few explicitly referred to histopathological findings, despite histopathology being considered the diagnostic gold standard for acute appendicitis.

Regarding surgical technique, the information reported across studies is highly limited and heterogeneous. Some authors mentioned at some point in the manuscript that a laparoscopic approach was used [20,23], but did not specify whether any conversions occurred or whether 100% of patients underwent laparoscopy. Other studies included only laparoscopic procedures and explicitly excluded open appendectomies or conversions [24,31,33]. Authors using national databases adjusted odds ratios for the “laparoscopic technique” variable and reported the proportion of patients undergoing laparoscopy, with significant differences between groups (p < 0.01) [21]. In one study, propensity score matching was used to balance the number of open and laparoscopic procedures between groups, while retaining both techniques in the analysis [30]. In contrast, some authors provided no details on surgical technique [22,28,32], while others included both open and laparoscopic appendectomies indiscriminately [25,27]. One study explicitly stated that the intraoperative technique was not standardized [26]. Finally, another study specified that all procedures were laparoscopic, although one conversion occurred in the NHA group and was not excluded [29].

### Post-Discharge Oral Home Antibiotics vs. No-Home Antibiotics

The intervention group (IG) was defined as the NHA group or as those managed under a protocol specifically implemented to reduce or eliminate post-discharge antibiotic use. In most studies, this corresponded to a “new protocol” cohort explicitly designed to minimize or avoid OHA. In some cases, raw data were reported specifically for NHA patients; in others, outcomes reflected the entire cohort managed under the revised protocol, even if not all patients were strictly NHA. The comparator group (CG) included patients who either received OHA at discharge or were managed according to standard protocols that did not restrict post-discharge antibiotic use. However, these protocols did not always guarantee that all patients in the CG actually received OHA. We clearly specified in the manuscript whether raw data were available or not for each study, and analyses were conducted accordingly.

A total of six studies directly compared patients discharged with OHA versus those discharged without them (NHA) [21,27,28–30,33], while eight studies compared outcomes across two distinct clinical protocols—typically a pre-implementation (baseline) phase versus a post-implementation phase [20,22–26,31,32]. In most cases, the pre-protocol group reflected routine or institutionally guided OHA administration, whereas the post-protocol group incorporated a clinical modification aimed at reducing OHA use. Although these constituted targeted interventions, they were primarily implemented as part of institutional practice changes rather than formal research initiatives, and most studies retained an observational retrospective design.

The specific characteristics of these institutional protocols, including their rationale and implications for antibiotic stewardship and discharge practices, are detailed in Table 1. Notably, one study (Ferguson [24]) inverted this structure: the authors standardized the routine prescription of OHA and designated that cohort as the intervention group. For this review, we reclassified this group as the comparator (CG) to maintain consistency in operational definitions across studies. These criteria and their application are detailed in Table 1.

Table 1 summarizes the data extracted from the fourteen studies that evaluated the role of post-discharge OHA in CAA.

### Risk of Bias Assessment

In the domain of bias due to confounding, seven studies were rated as having a serious risk of bias [20,21,24,25,27,29,33], while the remaining seven were rated as moderate [22,23,26,28,30,31,32]. None were considered at low risk. One of the main reasons for assigning a serious risk rating was the presence of significant differences in baseline patient characteristics, with systematically sicker patients more likely to receive OHA. For instance, in the study by Anderson et al. [21], patients in the OHA group had significantly higher preoperative leukocyte counts, a greater prevalence of systemic inflammatory response syndrome, and higher rates of predischarge SSI. This phenomenon—known as *confounding by indication*—precludes a clear distinction between the effects of the treatment and the underlying severity of illness. Another key reason was the presence of concurrent interventions; for example, the study by Ferguson et al. [24] coincided with a quality improvement initiative on the use of intraoperative abdominal drains and a separate pilot randomized trial of intra-abdominal irrigation with povidone-iodine, making it difficult to isolate the effect of the antibiotic protocol alone. Other reasons included the use of historical controls, which introduces temporal confounding, and massive, unadjusted baseline differences in disease severity between hospital cohorts being compared (such as in the case of Desai et al.) [20].

For bias in the selection of participants into the study, no studies were identified as having a serious risk, 5 were rated as moderate [20,24,28,30,31], and 9 were rated as low risk [21–23,25–27,29,32,33].

In the domain of bias in the classification of interventions, no studies were found to pose a serious risk, 7 were rated as moderate [21,23,25,27,28,30,33], and 7 were rated as low risk [20,22,24,26,29,31,32].

Regarding bias due to deviations from intended interventions, one study was rated as having a serious risk [23], nine as moderate [21,24,25,27,28–32], and four as low risk [20,22,26,33]. The serious risk rating was assigned because the study explicitly reported that "overall compliance with protocol guidelines was not ideal," with participation at the surgeon’s discretion. This led to substantial crossover, in which patients in the new protocol group continued receiving older treatments, contaminating the comparison groups and biasing the results toward finding no difference.

Regarding bias due to missing data, no studies posed a serious risk; 8 were rated as moderate [20,22,23,25–27,29,33]; and six were rated as low risk [21,24,28,30–32].

Regarding bias in the measurement of outcomes, none of the studies posed a serious risk, one study posed a moderate risk [20], and 13 were rated as low risk [21–33].

Finally, in the domain of bias in the selection of the reported result, no study was judged to be at serious or moderate risk; all 14 studies were rated as having a low risk of bias [20–33].

Figure 2 visually displays the ROBINS-I risk-of-bias assessments across individual domains for each included study.

**Figure 2.**
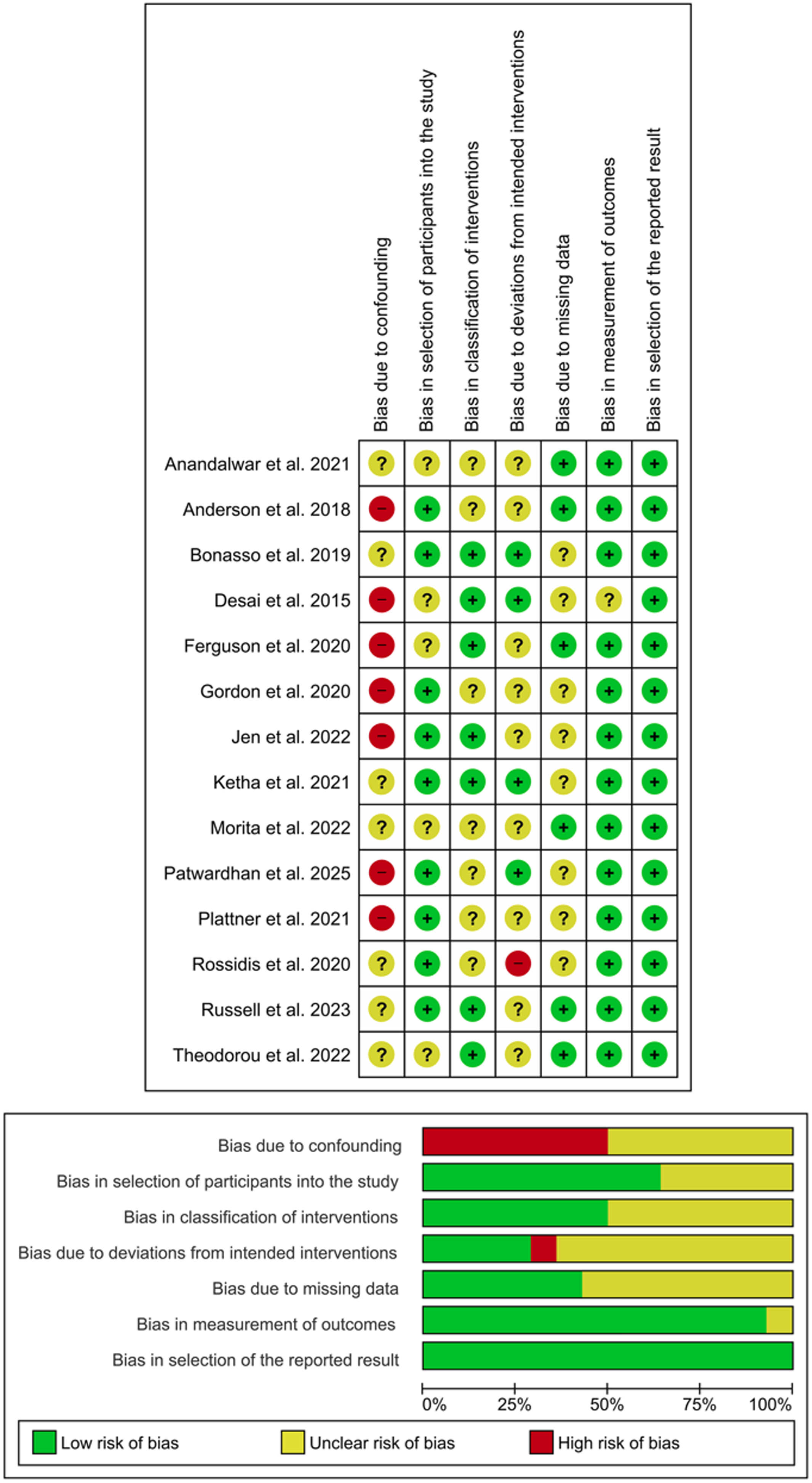
Graphical summary of the risk of bias assessment using the ROBINS-I tool for the included non-randomized studies. Green circles with a plus sign indicate low risk of bias, yellow circles with a question mark represent moderate risk, and red circles with a minus sign denote high risk of bias.

Supplementary File 4 and Table 1 provide detailed information on the data used for each study, including whether raw exposure data were available.

### Intra-abdominal Abscess (Oral Home Antibiotics vs. No Home Antibiotics)

Six studies reported IAA rates stratified by treatment group [20,22,24,25,26,27]. Bonasso et al. [22] presented two separate cohorts. Five cohorts found no significant differences in IAA rates between groups [20,22,25,26,27]. One study reported a significantly higher IAA rate in the IG than in the CG [24]. In contrast, one cohort reported a significantly higher IAA rate in the CG than in the IG [22].

Notably, the only study reporting increased IAA rates in the IG was Ferguson et al. [24], which uniquely implemented a protocol to standardize the routine use of OHA, rather than to reduce them. This contrasts with the rest of the literature, where interventions consistently aimed to minimize post-discharge antibiotic use.

### Meta-Analysis for Intra-abdominal Abscess (Oral Home Antibiotics vs. No Home Antibiotics).

For the IAA meta-analysis, a random-effects model was fitted using restricted maximum likelihood (REML) with truncated modified Knapp–Hartung (mKH) adjustments. Seven studies were included, encompassing 688 patients in the IG and 1,011 patients in the CG. This meta-analysis included a combination of studies reporting raw group-level data (OHA vs NHA) and others presenting outcomes for cohorts managed under pre- or post-intervention protocols. The pooled risk ratio (RR) comparing IG versus CG was 1.23 (95% CI: 0.62-2.45; model p = 0.49), indicating no significant difference in IAA rates between the groups. Between-study heterogeneity was low (I² = 25.87%; Cochran’s Q p = 0.05), and the estimated between-study variance was τ² = 0.08. The 95% PI ranged from 0.441 to 3.441. Leave-one-out sensitivity analysis revealed that excluding any single study did not materially alter the overall result or achieve statistical significance in any iteration, supporting the robustness of the null finding (Figure 3). A sensitivity analysis was conducted on studies reporting raw event data for both OHA and NHA [20,22,25,27]. This analysis included five studies and, under a REML random-effects model with mKH adjustment, yielded a pooled RR of 0.94 (95% CI 0.23–3.86), τ² = 0.48, I² = 75%, Q = 9.71 (p = 0.05), with the model test non-significant (p = 0.62). Given that fewer than ten studies were included, between-study heterogeneity was not further explored using funnel plots or formal statistical tests for small-study effects (e.g., Egger’s or Begg’s tests). Similarly, no meta-regression analyses were conducted due to the low number of studies and limited statistical power.

**Figure 3.**
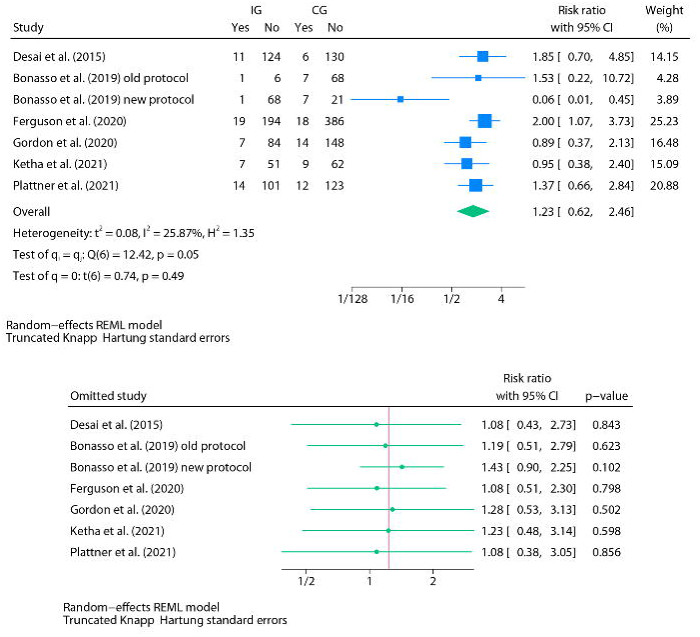
<u>Top</u>: Forest plot of the random-effects meta-analysis for intra-abdominal abscesses (IAA) using REML with Knapp–Hartung adjustment for confidence intervals (IG vs. CG). <u>Bottom</u>: Leave-one-out forest plot for the same analysis.

For IAA, the certainty of evidence was rated very low after downgrading for serious risk of bias, serious inconsistency, and serious imprecision, with no applicable upgrading domains.

### Surgical Site Infection (Oral Home Antibiotics vs. No Home Antibiotics)

Seven studies reported SSI rates stratified by treatment group [21,23,24,29,31–33]. Five studies found no significant differences in SSI rates between groups [23,24,29,32,33]. One study reported a significantly higher SSI rate in the IG than in the CG [31], whereas another reported a significantly higher SSI rate in the CG than in the IG [21].

### Meta-Analysis for Surgical Site Infection (Oral Home Antibiotics vs. No Home Antibiotics)

For the SSI meta-analysis, a random-effects model was fitted using REML. Seven studies were included, encompassing 3,858 patients in the IG and 4,610 in the CG. This meta-analysis included a combination of studies reporting raw group-level data (OHA vs NHA) and others presenting outcomes for cohorts managed under pre- or post-intervention protocols. The pooled RR comparing IG versus CG was 0.77 (95% CI: 0.61-0.96; model p = 0.02), indicating a significant difference in SSI rates. Between-study heterogeneity was negligible (I² = 0%; Cochran’s Q p = 0.55), and the estimated between-study variance was τ² = 0. The 95% PI ranged from 0.571 to 1.033. Leave-one-out sensitivity analysis showed that this association was not robust, as excluding Anderson et al. yielded a non-significant effect (RR = 0.96; 95% CI: 0.57– 1.61; p = 0.88) (Figure 4). In the sensitivity analysis restricted to studies reporting crude patient-level exposure data (n=2), the pooled point estimate shifted in the opposite (reversed) direction (RR 2.34; 95% CI 0.72–7.64; p = 0.16), suggesting higher SSI rates in the OHA group when the analysis is based on true exposure rather than protocol-defined cohorts. This reversal of direction indicates that protocol-based rather than true exposure-level comparisons drive the apparent benefit of NHA and do not reflect a consistent protective effect. Given that fewer than ten studies were included, between-study heterogeneity was not further explored using funnel plots or formal statistical tests for small-study effects (e.g., Egger’s or Begg’s tests). Similarly, no meta-regression analyses were conducted due to the low number of studies and limited statistical power.

**Figure 4.**
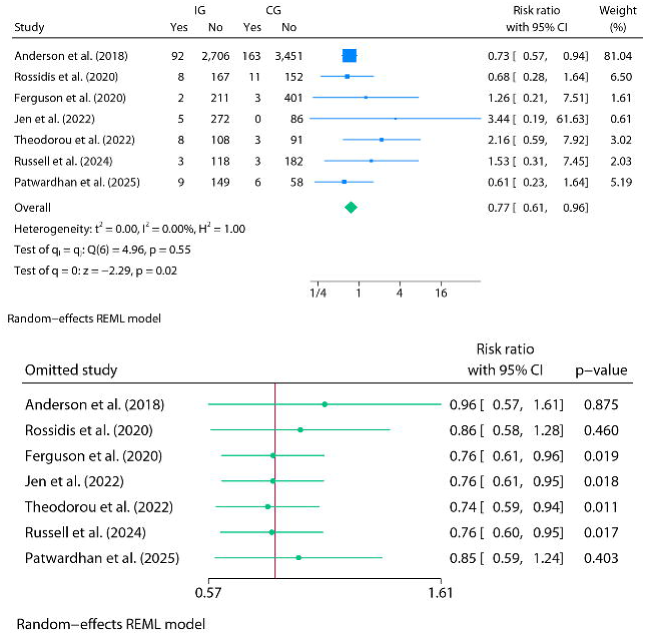
<u>Top</u>: Forest plot of the random-effects meta-analysis for surgical site infections (SSI) using REML (IG vs. CG). <u>Bottom</u>: Leave-one-out forest plot for the same analysis.

For SSI, the certainty of evidence was rated very low after downgrading for serious risk of bias and serious imprecision, with no applicable upgrading domains.

### Organ Space Infection (Oral Home Antibiotics vs. No Home Antibiotics)

Three studies reported OSI rates stratified by treatment group [28,29,32]. None of them found significant differences in OSI rates between groups [28,29,32].

### Meta-Analysis for Organ Space Infection (Oral Home Antibiotics vs. No Home Antibiotics)

For the OSI meta-analysis, a random-effects model was fitted using REML. Three studies were included, encompassing 685 patients in the IG and 558 in the CG. In the case of Anandalwar et al., only data from the propensity-matched cohort were included to avoid duplicating data from the same population and to ensure study independence. This meta-analysis included a combination of studies reporting raw group-level data (OHA vs NHA) and others presenting outcomes for cohorts managed under pre- or post-intervention protocols. The pooled RR comparing IG versus CG was 1.19 (95% CI: 0.73-1.93; model p = 0.48), indicating no significant difference in OSI rates. Between-study heterogeneity was negligible (I² = 0%; Cochran’s Q p = 0.47), and the estimated between-study variance was τ² = 0. Accordingly, confidence intervals were calculated using conventional Wald-type methods, as recommended when no heterogeneity is detected. The 95% PI ranged from 0.052 to 27.487. Leave-one-out sensitivity analysis revealed that no single study significantly altered the overall result or achieved statistical significance in any iteration (Figure 5). A sensitivity analysis was conducted on studies reporting raw event data for both OHA and NHA. This analysis included two studies [28,29] and, under a REML random-effects model without CI adjustment, yielded a pooled RR of 1.20 (95% CI 0.56–2.56; p = 0.64), with τ² = 0.10, I² = 29.08%, and Q = 1.41 (p = 0.24).

**Figure 5.**
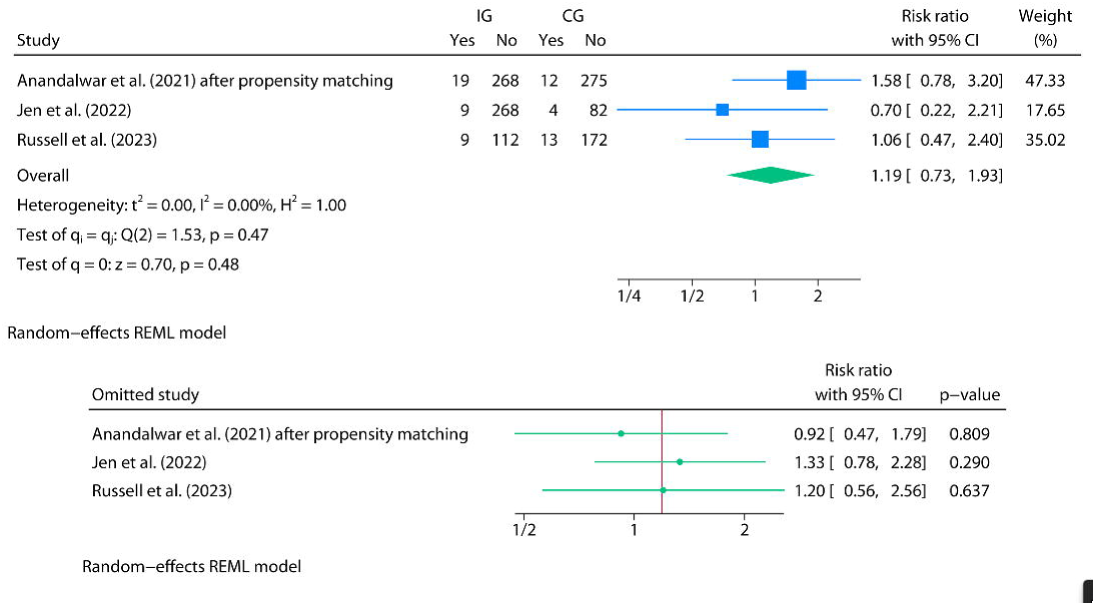
<u>Top</u>: Forest plot of the random-effects meta-analysis for organ/space infections (OSI) using REML (IG vs. CG). <u>Bottom</u>: Leave-one-out forest plot for the same analysis.

Using a REML model with mHK produced an extremely wide estimate (RR 1.20, 95% CI 0.09– 164.24; p = 0.72). These results illustrate that meta-analyses based on only two heterogeneous studies are methodologically unstable: Wald-type CIs tend to be overly optimistic, whereas mHK/HKSJ adjustments often become overly conservative under such conditions. Given that fewer than ten studies were included, between-study heterogeneity was not further explored using funnel plots or formal statistical tests for small-study effects (e.g., Egger’s or Begg’s tests). Similarly, no meta-regression analyses were conducted due to the low number of studies and limited statistical power.

For OSI, the certainty of evidence was rated very low after downgrading for serious risk of bias and serious imprecision, with no applicable upgrading domains.

### Readmissions (Oral Home Antibiotics vs. No Home Antibiotics)

Thirteen studies reported RA rates stratified by treatment group [21–33]. Ten studies found no significant differences in RA rates between groups [22–29, 31, 32]. One study reported a significantly higher RA rate in the IG compared to the CG [24], whereas two studies reported significantly higher RA rates in the CG than in the IG [21, 30]. In the case of Patwardhan et al. [33], although the difference did not reach statistical significance, a marginal p-value (0.06) was observed, suggesting a higher rate of events in the IG. Although most studies defined RA as occurring within 30 days of initial discharge, some—such as Morita et al. [30]—used a 60-day timeframe, which may influence effect estimates due to the extended follow-up window.

### Meta-Analysis for Readmissions (Oral Home Antibiotics vs. No Home Antibiotics)

For the meta-analysis of RA, a random-effects model was fitted using REML with mKH adjustments. Bonasso et al. [22], which also reported RA rates, was excluded from the pooled analysis due to internal inconsistencies in patient numbers and reported events elsewhere in the study, precluding reliable extraction for pooling. Twelve studies were included, encompassing 7,159 patients in the IG and 8,053 in the CG. In the cases of Anandalwar et al. (2021) and Morita et al. (2022) [28,30], only data from the propensity-matched cohorts were included to ensure independence of observations. This meta-analysis included a combination of studies reporting raw group-level data (OHA vs NHA) and others presenting outcomes for cohorts managed under pre- or post-intervention protocols. The pooled RR comparing IG versus CG was 1.07 (95% CI: 0.78-1.44; model p = 0.66), indicating no significant difference in readmission rates. Between-study heterogeneity was substantial (I² = 64.97%; Cochran’s Q p = 0.00), with a between-study variance of τ² = 0.11. The 95% PI ranged from 0.475 to 2.394. Because this was the only meta-analysis including ≥10 studies, small-study effects and publication bias were formally assessed. Leave-one-out sensitivity analysis revealed that no single study materially affected the overall result or achieved statistical significance. The exclusion of Ferguson et al. (2020) produced the most pronounced shift, but the result remained non-significant (Figure 6). Visual inspection of the contour-enhanced funnel and Galbraith (radial) plots did not reveal asymmetry, which was consistent with non-significant Egger and Begg tests (Egger p = 0.18; Begg p = 0.19). A L’Abbé plot was generated exclusively to illustrate between-study variability (heterogeneity) (Figure 7).

**Figure 6.**
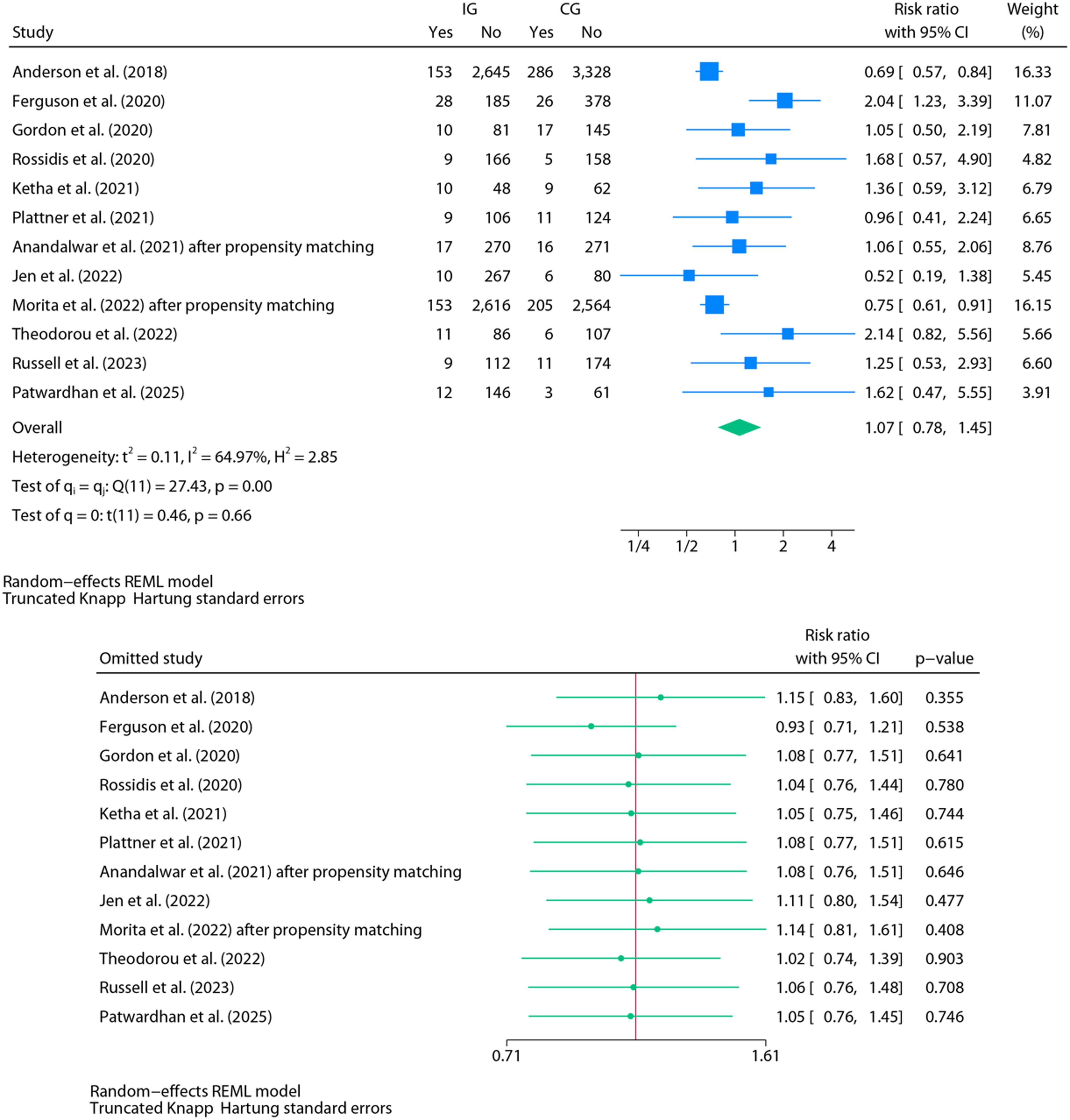
<u>Top</u>: Forest plot of the random-effects meta-analysis for hospital readmissions (RA) using REML with Knapp–Hartung adjustment (IG vs. CG). <u>Bottom</u>: Leave-one-out forest plot for the same analysis.

**Figure 7.**
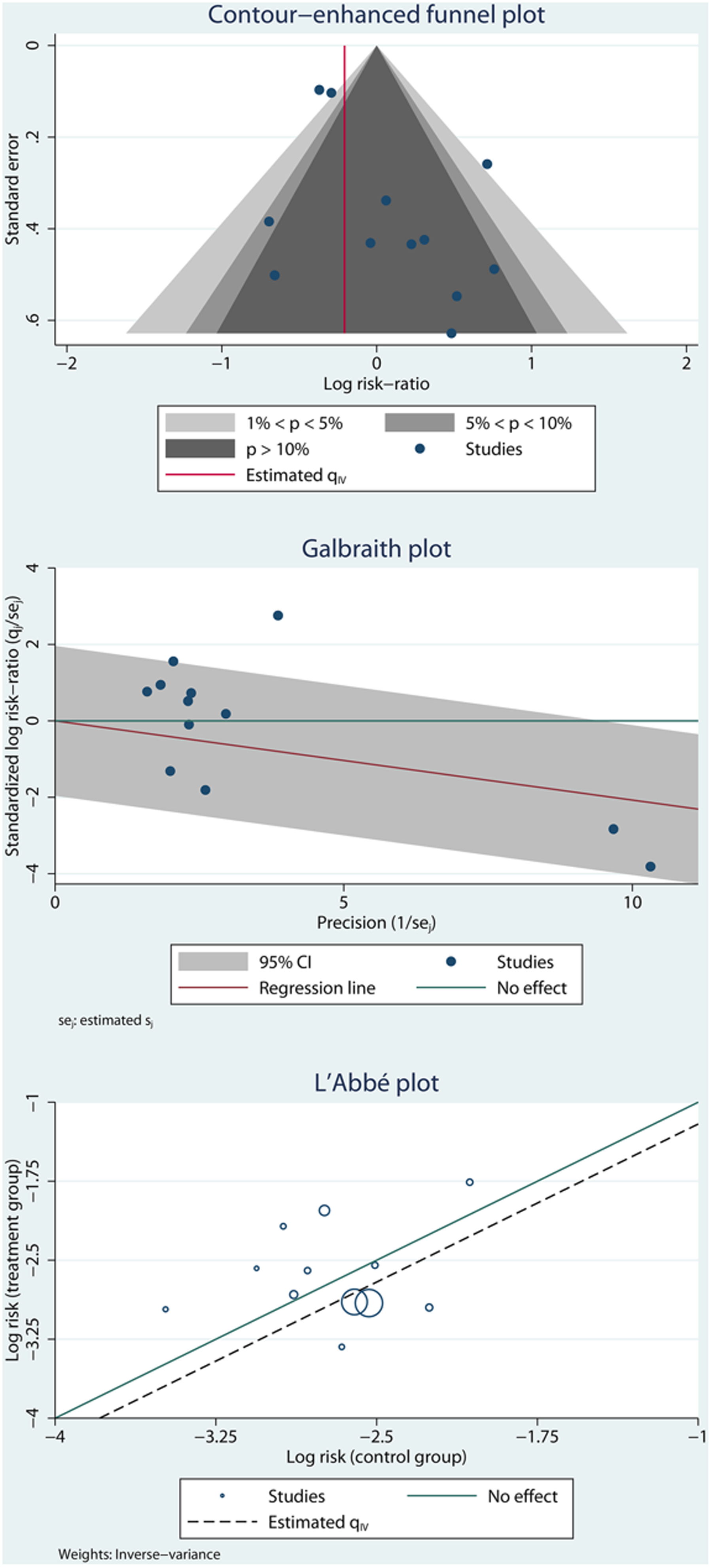
<u>Top</u>: contour-enhanced funnel plot assessing small-study effects and potential publication bias. <u>Middle</u>: Galbraith (radial) plot assessing overall heterogeneity and study precision. <u>Bottom</u>: L’Abbé plot illustrating between-study variability in event rates (heterogeneity exploration).

A complementary sensitivity analysis, including the five studies with available raw event data, was also performed. Under a REML random-effects model with mHK adjustment, the pooled effect was RR 0.78 (95% CI 0.61–1.01; p = 0.05), with τ² = 0.00, I² = 0%, and Q = 2.53 (p = 0.55), indicating no detectable heterogeneity. This analysis of patient-level exposure data suggests that OHA may be associated with an increased risk of RA. Protocol-based comparisons are methodologically weaker because they conflate exposure with institutional timing and practice evolution. By contrast, crude exposure studies provide a cleaner estimate at the patient level. The fact that the association becomes stronger and statistically significant once protocol contamination is removed suggests that protocol-based designs were masking a potential adverse signal.

An univariable meta-regression was conducted, using differences in predischarge total leukocyte count (TLC) as a categorical moderator to examine whether discharge inflammatory status influenced the association between OHA and the risk of RA. The analysis compared studies reporting no differences in TLC between groups (reference category) with those reporting significant differences—most often reflecting higher TLC levels in the comparator group (CG, i.e., OHA)—and studies that did not specify this information. The analysis found that this variable was not significantly associated with the pooled risk ratios (β = –0.11; p = 0.66) and did not explain any of the observed between-study heterogeneity (R² = 0.00%). While it is plausible that these differences reflect clinical decision-making, in which patients with signs of residual inflammation were more likely to receive antibiotics, this meta-regression found no statistical evidence that this factor influenced the overall outcome. However, as the category with significant TLC differences included only two studies, this trend should be interpreted with caution, given the potential influence of individual study effects and limited statistical power.

Lastly, a univariable meta-regression comparing high-versus moderate-risk studies showed no evidence of effect modification (logRR difference: –0.20; 95% CI: –0.78 to 0.38; p = 0.50), with R² = 0%. This indicates that the risk of bias does not account for the observed variability and offers no explanatory value for the treatment effect.

For RA, the certainty of evidence was rated very low after downgrading for serious risk of bias, serious inconsistency, and serious imprecision, with no applicable upgrading domains.

### Secondary Outcomes (Oral Home Antibiotics vs. No Home Antibiotics)

Some studies also examined alternative outcomes such as composite post-discharge morbidity [21], reoperations (RO) [21,23,24], and Emergency Department visits (EDV) [21,23,24,29]. Detailed outcome data are available in Table 1. Given the low number of studies reporting these outcomes and the heterogeneity and dispersion of available data, no meta-analytic models were fitted for these endpoints.

## Discussion

This systematic review and meta-analysis provide the most comprehensive synthesis to date of the impact of OHA following surgical management of CAA in children. Across multiple outcomes—including IAA and OSI—no consistent benefit was observed with OHA use. Indeed, SSI were significantly more frequent among patients in the CG in the primary model; however, this finding was not robust and disappeared once analyses were restricted to crude exposure data, where the point estimate shifted in the opposite direction. This pattern is consistent with confounding by indication in protocol-based cohorts. Therefore, the primary SSI signal should be interpreted as artefactual rather than indicative of benefit. Lastly, in crude exposure analyses, RA rates were lower in the NHA cohort. Both individual study estimates and pooled risk ratios failed to support routine administration of post-discharge OHA. Despite their widespread use, this is the first synthesis to assess their effectiveness across these core outcomes. Notably, several analyses showed a trend in favor of NHA discharge strategies for RA, and in the exposure-restricted subset, this difference reached statistical significance, consistent with a potential harmful signal rather than a neutral effect.

Our findings challenge the traditional rationale for extended antibiotic coverage after discharge in pediatric CAA, particularly in cases with adequate source control. Previous guidelines and institutional practices often endorsed OHA as a safeguard against latent infections or undetected abscess formation. However, the current evidence, including recent protocol-based studies aimed at restricting OHA, suggests that this approach may offer no additional benefit—and may contribute to overtreatment, antimicrobial resistance, and unnecessary drug exposure. This underscores the importance of aligning postoperative management with principles of antimicrobial stewardship, especially in pediatric patients.

Interestingly, the only study reporting a significantly higher RA and IAA rate in the NHA group [24] was also the only one to implement a protocol aimed at standardizing rather than limiting post-discharge OHA. This divergence contrasts with the broader trend in literature, where interventions were designed to reduce or eliminate OHA use. Thus, this deviation highlights how institutional culture and protocol framing may shape not only practices but also outcomes. It also raises the possibility of confounding by protocol context—underscoring the need for randomized comparisons within standardized care frameworks.

From a clinical standpoint, several included studies explored risk-adapted prescribing by preferentially giving OHA to patients with higher inflammatory markers or more severe intraoperative findings. However, because these decisions were observational and confounded by indication, the certainty of evidence supporting this approach is very low under GRADE. Therefore, while the concept may be clinically appealing, the current data do not validate risk-adapted selection of OHA, and any such strategy remains unproven until tested in higher-certainty designs. Although some relevant RCTs, such as the APPIC trial [34], evaluated inpatient intravenous antibiotic duration rather than post-discharge regimens, they provide indirect contextual relevance: even earlier in the treatment pathway, shortening antibiotic exposure has not resulted in worse outcomes when adequate source control is achieved. Its applicability to OHA is therefore indirect, but the directionality is consistent — the burden of justification seems to lie on prolonged antibiotic exposure, not on reduction.

Another potential source of bias stems from clinical decision-making criteria for prescribing antibiotics at discharge. Although some studies reported predischarge TLC and their comparability between groups, this information was frequently absent or insufficiently detailed. This limits the ability to determine whether the inflammatory status at discharge systematically influenced treatment allocation. In our exploratory meta-regression, studies reporting statistically significant differences in TLC between groups—typically indicating higher values in the antibiotic group—showed a trend toward lower pooled RR in the NHA. While this finding should be interpreted with caution due to the limited power and potential confounding, it suggests that patients perceived as being at higher risk (e.g., those with elevated TLC) may have been selectively prescribed antibiotics yet did not show a corresponding reduction in readmission rates. This paradoxical trend may reflect residual confounding and highlight the complexity of interpreting observational data. This hypothesis-generating observation highlights the need for future studies to document clinical parameters that inform discharge decisions rigorously and to control for these factors in comparative analyses. Structured discharge algorithms incorporating inflammatory markers may help reduce selection bias and improve comparability in future research.

Importantly, confounding by indication should not be interpreted merely as a limitation but as a central interpretive lens. The bias is directional, not random: children perceived as higher-risk (e.g., elevated predischarge TLC or systemic inflammation) were systematically more likely to receive OHA. If OHA were truly protective, this clinical enrichment of the treated cohort should have amplified its observable benefit. Instead, no benefit was seen, and when analyses were restricted to studies with individual-level exposure data (OHA vs NHA), the signal moved toward potential harm. Far from weakening the null finding, this strengthens it: a therapy that fails to demonstrate benefit even in the subgroup most selectively “enriched” to benefit is unlikely to be effective in routine practice.

Another important consideration is the limited reporting and control of variables that may substantially influence outcomes but were inconsistently addressed across studies. For example, none of the included articles provided objective measures of patient compliance with prescribed oral antibiotics—a critical determinant of treatment efficacy. Similarly, the relationship between discharge antibiotic strategy and surgical technique (e.g., laparoscopic versus open approach), intraoperative findings, or postoperative management protocols was rarely explored in depth. These factors could interact with the effect of OHA and confound outcome estimates. Additionally, key elements such as discharge timing, the presence of drains, or concomitant antibiotic regimens (e.g., IV continuation) were often insufficiently detailed. Potential differences in microbiological etiology or resistance patterns were also largely unaddressed, limiting biological interpretability and external applicability. These gaps highlight the need for future prospective studies to standardize and report such variables systematically, enabling more precise estimation of the clinical utility—and limitations—of routine OHA in CAA.

From a biological standpoint, the rationale for OHA in this setting remains biologically questionable. The peritoneal cavity, particularly in the context of localized postoperative inflammation and fibrinous exudate, may offer limited penetration for orally administered antibiotics—especially in the absence of systemic signs of infection. Furthermore, the oral route introduces variability in absorption, particularly in children with reduced appetite, postoperative ileus, or gastrointestinal dysmotility. These pharmacokinetic limitations, combined with incomplete data on compliance and dosing, raise concerns about the real-world efficacy of oral agents in preventing deep or loculated infections. Additionally, unnecessary antibiotic exposure may select for resistant microbial strains, potentially altering the child’s microbiome and increasing future vulnerability to infection. Altogether, these pharmacological considerations further support a risk-adapted rather than uniform approach to OHA use.

Moreover, none of the included studies systematically reported adverse events associated with oral antibiotic use, such as diarrhea, candidiasis, or intestinal dysbiosis, which represents an important limitation. The absence of statistically significant differences should not be interpreted as evidence of equivalence; limited power and unmeasured non-adherence may have attenuated the observed effects. However, the consistent directional pattern across leave-one-out and exposure-restricted analyses suggests that the null findings are unlikely to be explained solely by insufficient power. This omission prevents quantification of the potential collateral harms of unnecessary antibiotic administration, but it indirectly reinforces one of the core principles of antibiotic stewardship: avoiding treatments that provide no clinical benefit and may instead cause preventable adverse effects. In this context, our findings are particularly applicable to patients with favorable postoperative evolution—namely, those who are afebrile, tolerating oral intake, in good general condition, and with normalized inflammatory markers at discharge—for whom continued antibiotic treatment beyond discharge may offer no additional benefit. It should also be emphasized that the majority of included studies administered intravenous antibiotics during hospitalization, and thus, these results should not be extrapolated to settings in which early discharge with oral antibiotics alone is considered—an approach not evaluated in this review. Avoiding unnecessary OHA in these patients may also reduce the risk of avoidable harm in an already recovering pediatric population.

Lastly, an essential methodological concern highlighted by this review is the substantial variability in the reporting of surgical technique. We observed marked differences in both the inclusion/exclusion criteria based on surgical approach (open vs. laparoscopic) and the overall quality of reporting. While some studies explicitly excluded open appendectomies or documented conversions, others merely stated that laparoscopy was used without specifying whether this applied to all patients or whether any conversions occurred. Several studies failed to report the surgical approach altogether. This lack of standardization is concerning, given well-established evidence that open appendectomies carry a higher risk of surgical site infection than laparoscopic procedures, potentially introducing uncontrolled confounding into the interpretation of results. Similarly, relevant operative variables such as surgical duration—which may reflect case complexity and is associated with postoperative outcomes—were scarcely reported across the included studies. Given the well-established association between surgical approach and postoperative infection risk, this lack of standardization represents a critical gap in current evidence. Future research should systematically collect and transparently report key surgical variables to enable more accurate risk adjustment and a more robust interpretation of findings.

This review has several strengths. It adheres to PRISMA and Cochrane methodological standards. It applies modern, rigorous statistical techniques, such as random-effects REML meta-analysis with robust confidence intervals (HKSJ and the modified Knapp–Hartung) and prediction intervals, leave-one-out sensitivity analyses, and, when feasible, assessment of publication bias. Univariable meta-regression was used to explore sources of heterogeneity, including differences in predischarge TLC and the type of comparator group (raw OHA/NHA vs protocol-defined). Particular care was taken to distinguish studies reporting individual-level data from those reporting outcomes based on pre- and post-protocol cohorts, and all analyses were conducted accordingly. Importantly, statistical results were interpreted in light of the clinical context, considering how discharge practices and patient selection may have influenced both antibiotic use and outcomes. However, relevant limitations must be acknowledged. As detailed in our Results, a primary limitation is the substantial variability in the definition of CAA across studies. Definitions spanned a spectrum of findings (e.g., visible perforation, abscess, free fecalith) based on intraoperative, histopathological, or even unspecified criteria. This lack of standardization complicates direct comparisons and is a significant source of heterogeneity. Furthermore, individual-level OHA and NHA raw data were also often unavailable. Many studies lacked control for key variables such as antibiotic type, dosage, or precise discharge criteria. Additionally, the complications—particularly IAA and OSI—were not extensively characterized across articles, e.g., omitting severity grading or volumetric analysis of abscesses. Most studies were retrospective and based in the USA, and those using large databases, while offering substantial sample sizes, often lacked granular clinical information. Notably, only one study [27] included solid microbiological data, limiting the biological interpretability of the outcomes. Accordingly, the certainty of evidence was rated as low or very low across all primary outcomes using the GRADE framework [35], primarily due to the observational nature of the studies, moderate to high heterogeneity in some analyses, and imprecision of effect estimates.

These conclusions should be interpreted with caution, given the overall low certainty of evidence. Lastly, because several included studies used NSQIP-derived cohorts, partial patient overlap cannot be definitively excluded. Individual-level identifiers are not available in NSQIP publications, preventing formal de-duplication, and therefore, a residual risk of double-counting remains. Although we found no demonstrable overlap based on study periods and sampling frames, this possibility should be acknowledged when interpreting pooled estimates.

In light of these findings, the routine prescription of OHA after discharge in pediatric patients with surgically treated CAA appears unwarranted. Across 26,000 patients, no consistent benefit was observed for IAA, SSI, OSI, or RA. Importantly, subgroup crude analyses even suggest that OHA may increase the risk of readmission, particularly in patients selected for treatment based on residual inflammation (TLC). However, the current body of evidence is based exclusively on observational studies, with heterogeneous definitions, limited adjustment for confounders, and no data on patient adherence. While the convergent pattern of results strengthens the biological and methodological plausibility of no benefit, the GRADE rating of very low certainty means that inference must remain cautious. The present synthesis supports a presumption *against* routine OHA, but clinical equipoise persists, and only well-designed prospective trials can definitively settle this question.

There is an urgent need for adequately powered, multicenter randomized controlled trials that rigorously control for clinical, surgical, and microbiological variables to definitively establish the role—or lack thereof—of oral home antibiotics in this setting. Future studies should clearly specify the surgical technique employed (e.g., open vs laparoscopic, use of peritoneal lavage), characterize microbiological findings and antibiotic susceptibility profiles, and distinguish individual-level antibiotic exposure (OHA vs NHA) rather than relying solely on protocol-based groupings. Critically, future trials must be based on a clear, internationally accepted consensus definition of CAA. Additionally, consistent definitions for outcomes such as IAA, SSI, and readmissions are essential to ensure comparability across studies. Until such trials are available, a cautious, individualized approach to OHA prescription—grounded in objective resolution criteria—may best balance safety with antimicrobial stewardship in pediatric CAA.

## Data Availability

All data used for the meta-analytical models are available in the accompanying supplementary dataset file.

## CRediT authorship contribution statement

**JAM:** Conceptualization and study design; literature search and selection; data curation and extraction; formal analysis; investigation; methodology; project administration; resources; validation; visualization; writing – original draft; writing – review and editing.

**MRJ:** Literature search and selection; data curation and extraction; writing, review, and editing.

## ETHICAL APPROVAL

This study did not involve the participation of human or animal subjects, and therefore, IRB approval was not sought.

## Supplementary Materials

**Supplementary File 1.** PRISMA checklist

**Supplementary File 2.** Full search strategy.

**Supplementary File 3.** Inclusion and exclusion criteria.

**Supplementary File 4.** Full dataset

## References

[1]. Chionye R. Ossai, Lu Pu, David Kaelber; Using Aggregated Data from 1.4 Million Pediatric Patients to Describe the Epidemiology and Demographic Characteristics of Appendicitis. Pediatrics July 2020; 146 (1_MeetingAbstract): 232–234. 10.1542/peds.146.1MA3.232b

[2]. Omling E, Salö M, Saluja S, Bergbrant S, Olsson L, Persson A, Björk J, Hagander L. Nationwide study of appendicitis in children. Br J Surg. 2019 Nov;106(12):1623–1631. doi: 10.1002/bjs.11298. Epub 2019 Aug 6. PMID: 31386195; PMCID: PMC6852580.

[3]. Georgeades C, Bodnar C, Bergner C, Van Arendonk KJ. Association of complicated appendicitis with geographic and socioeconomic measures in children. Surgery. 2024 Nov;176(5):1475–1484. doi: 10.1016/j.surg.2024.07.044. Epub 2024 Sep 3. PMID: 39232975.

[4]. Michelson KA, Bucher BT, Neuman MI. Cost and Late Hospital Care of Publicly Insured Children After Appendectomy. J Surg Res. 2024 May;297:41–46. doi: 10.1016/j.jss.2024.02.003. Epub 2024 Mar 1. PMID: 38430861; PMCID: PMC11023751.

[5]. Wang C, Li Y, Ji Y. Intravenous versus intravenous/oral antibiotics for perforated appendicitis in pediatric patients: a systematic review and meta-analysis. BMC Pediatr. 2019 Nov 4;19(1):407. doi: 10.1186/s12887-019-1799-6. PMID: 31684906; PMCID: PMC6827245.

[6]. Lam JY, Beaudry P, Simms BA, Brindle ME. Impact of implementing a fast-track protocol and standardized guideline for the management of pediatric appendicitis. Can J Surg. 2021 Jul 5;64(4):E364-E370. doi: 10.1503/cjs.005420. PMID: 34223740; PMCID: PMC8410463.

[7]. Muehlstedt SG, Pham TQ, Schmeling DJ. The management of pediatric appendicitis: a survey of North American Pediatric Surgeons. J Pediatr Surg. 2004 Jun;39(6):875–9; discussion 875-9. doi: 10.1016/j.jpedsurg.2004.02.035. PMID: 15185217.

[8]. Huttner B, Harbarth S, Carlet J, et al. Antibiotics are not automatic anymore—the French national campaign to cut antibiotic overuse. PLoS Med. 2010;7(7):e1000353. doi:10.1371/journal.pmed.1000353

[9]. Sabuncu E, David J, Bernède-Bauduin C, Pépin S, Leroy M, Boëlle PY, Watier L, Guillemot D. Significant reduction of antibiotic use in the community after a nationwide campaign in France, 2002-2007. PLoS Med. 2009 Jun 2;6(6):e1000084. doi: 10.1371/journal.pmed.1000084. Epub 2009 Jun 2. PMID: 19492093; PMCID: PMC2683932.

[10]. Page MJ, McKenzie JE, Bossuyt PM, Boutron I, Hoffmann TC, Mulrow CD, et al. The PRISMA 2020 statement: an updated guideline for reporting systematic reviews. BMJ. 2021 Mar 29;372:n71. doi: 10.1136/bmj.n71. PMID: 33782057; PMCID: PMC8005924.

[11]. Higgins JPT, Thomas J, Chandler J, Cumpston M, Li T, Page MJ, Welch VA (editors). Cochrane Handbook for Systematic Reviews of Interventions version 6.5 (updated August 2024). Cochrane, 2024. Available from www.cochrane.org/handbook.

[12]. Sterne JAC, Savović J, Page MJ, Elbers RG, Blencowe NS, Boutron I et al. RoB 2: a revised tool for assessing risk of bias in randomised trials. BMJ. 2019 Aug 28;366:l4898. doi: 10.1136/bmj.l4898. PMID: 31462531.

[13]. Sterne JA, Hernán MA, Reeves BC, Savović J, Berkman ND, Viswanathan M et al. ROBINS-I: a tool for assessing risk of bias in non-randomised studies of interventions. BMJ. 2016 Oct 12;355:i4919. doi: 10.1136/bmj.i4919. PMID: 27733354; PMCID: PMC5062054.

[14]. Veroniki AA, Jackson D, Viechtbauer W, Bender R, Bowden J, Knapp G, et al. Methods to estimate the between-study variance and its uncertainty in meta-analysis. Res Synth Methods. 2016;7(1):55–79. doi:10.1002/jrsm.1164

[15]. IntHout J, Ioannidis JP, Borm GF. The Hartung-Knapp-Sidik-Jonkman method for random effects meta-analysis is straightforward and considerably outperforms the standard DerSimonian-Laird method. BMC Med Res Methodol. 2014 Feb 18;14:25. doi: 10.1186/1471

[16]. Röver C, Knapp G, Friede T. Hartung-Knapp-Sidik-Jonkman approach and its modification for random-effects meta-analysis with few studies. BMC Med Res Methodol. 2015 Nov 14;15:99. doi: 10.1186/s12874-015-0091-1. PMID: 26573817; PMCID: PMC4647507

[17]. 2288-14-25. PMID: 24548571; PMCID: PMC4015721 Begg CB, Mazumdar M. Operating characteristics of a rank correlation test for publication bias. Biometrics. 1994;50(4):1088–1101.

[18]. Egger M, Davey Smith G, Schneider M, Minder C. Bias in meta-analysis detected by a simple, graphical test. BMJ. 1997;315(7109):629-634.

[19]. Shi L, Lin L. The trim-and-fill method for publication bias: practical guidelines and recommendations based on a large database of meta-analyses. Medicine (Baltimore). 2019 Jun;98(23):e15987. doi: 10.1097/MD.0000000000015987. PMID: 31169736; PMCID: PMC6571372.

[20]. Desai AA, Alemayehu H, Holcomb GW 3rd, St Peter SD. Safety of a new protocol decreasing antibiotic utilization after laparoscopic appendectomy for perforated appendicitis in children: A prospective observational study. J Pediatr Surg. 2015 Jun;50(6):912–4. doi: 10.1016/j.jpedsurg.2015.03.006. Epub 2015 Mar 14. PMID: 25812441.

[21]. Anderson KT, Bartz-Kurycki MA, Kawaguchi AL, Austin MT, Holzmann-Pazgal G, Kao LS, Lally KP, Tsao K. Home Antibiotics at Discharge for Pediatric Complicated Appendicitis: Friend or Foe? J Am Coll Surg. 2018 Aug;227(2):247–254. doi: 10.1016/j.jamcollsurg.2018.04.004. Epub 2018 Apr 20. PMID: 29680415.

[22]. Bonasso PC, Dassinger MS, Wyrick DL, Smith SD, Burford JM. Evaluation of white blood cell count at time of discharge is associated with limited oral antibiotic therapy in children with complicated appendicitis. Am J Surg. 2019 Jun;217(6):1099–1101. doi: 10.1016/j.amjsurg.2018.12.071. Epub 2019 Jan 3. PMID: 30639131.

[23]. Rossidis AC, Brown EG, Payton KJ, Mattei P. Implementation of an evidence-based protocol after appendectomy reduces unnecessary antibiotics. J Pediatr Surg. 2020 Nov;55(11):2379–2386. doi: 10.1016/j.jpedsurg.2020.07.001. Epub 2020 Jul 9. PMID: 32753275.

[24]. Ferguson DM, Parker TD, Arshad SA, Garcia EI, Hebballi NB, Tsao K. Standardized Discharge Antibiotics May Reduce Readmissions in Pediatric Perforated Appendicitis. J Surg Res. 2020 Nov;255:388–395. doi: 10.1016/j.jss.2020.05.086. Epub 2020 Jun 29. PMID: 32615311.

[25]. Gordon AJ, Choi JH, Ginsburg H, Kuenzler K, Fisher J, Tomita S. Oral Antibiotics and Abscess Formation After Appendectomy for Perforated Appendicitis in Children. J Surg Res. 2020 Dec;256:56–60. doi: 10.1016/j.jss.2020.05.082. Epub 2020 Jul 16. PMID: 32683057.

[26]. Ketha B, Stephenson KJ, Dassinger MS 3rd, Smith SD, Burford JM. Eliminating Use of Home Oral Antibiotics in Pediatric Complicated Appendicitis. J Surg Res. 2021 Jul;263:151–154. doi: 10.1016/j.jss.2020.12.059. Epub 2021 Feb 27. PMID: 33652177.

[27]. Plattner AS, Newland JG, Wallendorf MJ, Shakhsheer BA. Management and Microbiology of Perforated Appendicitis in Pediatric Patients: A 5-Year Retrospective Study. Infect Dis Ther. 2021 Dec;10(4):2247–2257. doi: 10.1007/s40121-021-00502-x. Epub 2021 Jul 21. PMID: 34287780; PMCID: PMC8572942.

[28]. Anandalwar SP, Graham DA, Kashtan MA, Hills-Dunlap JL, Rangel SJ. Influence of Oral Antibiotics Following Discharge on Organ Space Infections in Children With Complicated Appendicitis. Ann Surg. 2021 Apr 1;273(4):821–825. doi: 10.1097/SLA.0000000000003441. PMID: 31274648.

[29]. Jen J, Hwang R, Mattei P. Post-discharge antibiotics do not prevent intra-abdominal abscesses after appendectomy in children. J Pediatr Surg. 2023 Feb;58(2):258–262. doi: 10.1016/j.jpedsurg.2022.10.024. Epub 2022 Oct 29. PMID: 36428182.

[30]. Morita K, Fujiogi M, Michihata N, Matsui H, Fushimi K, Yasunaga H, Fujishiro J. Oral Antibiotics and Organ Space Infection after Appendectomy and Intravenous Antibiotics Therapy for Complicated Appendicitis in Children. Eur J Pediatr Surg. 2023 Feb;33(1):74–80. doi: 10.1055/a-1958-7915. Epub 2022 Oct 11. PMID: 36220134.

[31]. Theodorou CM, Lee SY, Lawrence Y, Saadai P, Hirose S, Brown EG. The Utility of Discharge Antibiotics in Pediatric Perforated Appendicitis Without Leukocytosis. J Surg Res. 2022 Jul;275:48–55. doi: 10.1016/j.jss.2022.01.024. Epub 2022 Feb 23. PMID: 35219251; PMCID: PMC10032146.

[32]. Russell KW, Skarda DE, Jones TW, Barnhart DC, Short SS. Cessation of Antibiotics for Complicated Appendicitis at Discharge Does Not Increase Risk of Post-operative Infection. J Pediatr Surg. 2024 Jan;59(1):91–95. doi: 10.1016/j.jpedsurg.2023.09.023. Epub 2023 Sep 26. PMID: 37858398.

[33]. Patwardhan UM, Kahan A, Eldredge RS, Russell KW, Lee J, Short SS, Padilla B, Cairo SB, Acker SN, Jensen AR, Kelley-Quon LI, Rothstein DH, Fialkowski EA, Chao SD, Gillory L, Pandya S, Diaz-Miron J, Ignacio RC Jr. Comparison of Postoperative Antibiotic Protocols for Pediatric Complicated Appendicitis: A Western Pediatric Surgery Research Consortium Study. J Pediatr Surg. 2025 Apr;60(4):162165. doi: 10.1016/j.jpedsurg.2025.162165. Epub 2025 Jan 9. PMID: 39827485.

[34]. de Wijkerslooth EML, Boerma EG, van Rossem CC, van Rosmalen J, Baeten CIM, Beverdam FH, Bosmans JWAM, Consten ECJ, Dekker JWT, Emous M, van Geloven AAW, Gijsen AF, Heijnen LA, Jairam AP, Melles DC, van der Ploeg APT, Steenvoorde P, Toorenvliet BR, Vermaas M, Wiering B, Wijnhoven BPL, van den Boom AL; APPIC Study Group. 2 days versus 5 days of postoperative antibiotics for complex appendicitis: a pragmatic, open-label, multicentre, non-inferiority randomised trial. Lancet. 2023 Feb 4;401(10374):366-376. doi: 10.1016/S0140-6736(22)02588-0. Epub 2023 Jan 17. PMID: 36669519.

[35]. Guyatt GH, Oxman AD, Vist GE, Kunz R, Falck-Ytter Y, Alonso-Coello P, Schünemann HJ; GRADE Working Group. GRADE: an emerging consensus on rating quality of evidence and strength of recommendations. BMJ. 2008 Apr 26;336(7650):924-6. doi: 10.1136/bmj.39489.470347.AD. PMID: 18436948; PMCID: PMC2335261.

