## Supplementary File 2 for "Post-discharge Oral Home Antibiotics Versus No Oral Home Antibiotics in Complicated Pediatric Appendicitis: A Systematic Review and Meta-analysis"

**Supplementary File 2: Search Strategy**

We will use a two-part search strategy to identify studies that meet the inclusion criteria and were published between inception and May 10th, 2025:

- We will search bibliographic databases in the fields of medicine and public health using a comprehensive search strategy targeting:
  - **Pediatric patients with surgically treated complicated acute appendicitis**,
  - **Oral antibiotic use at hospital discharge**, and
  - **Comparative studies evaluating post-discharge outcomes with and without oral antibiotic therapy**.
- We will also conduct forward and backward snowballing by screening the reference lists of all primary studies included in the review and identifying studies that have cited any of the included articles.

The following electronic databases will be searched: **PubMed, Web of Science, Scopus, Ovid,** **and** **Cochrane Central Register of Controlled Trials (CENTRAL)**.

**Search strategy**

**PubMed =** 57

("Appendicitis"[MeSH Terms] OR appendicitis[tiab]) AND

(complicat*[tiab] OR perforat*[tiab] OR gangren*[tiab]) AND

(pediatric*[tiab] OR child*[tiab] OR adolescent*[tiab] OR infant[tiab] OR "Pediatrics"[MeSH Terms]) AND (appendectomy[tiab] OR appendicectomy[tiab] OR surgery[tiab] OR "Appendectomy"[MeSH Terms]) AND ("Anti-Bacterial Agents"[MeSH Terms] OR antibiotic*[tiab] OR antimicrob*[tiab]) AND (oral[tiab] OR "post-discharge"[tiab] OR "at discharge"[tiab]) AND (random*[tiab] OR cohort[tiab] OR compar*[tiab] OR trial[tiab])

**WoS =** 106

TS=(appendicitis AND (complicat* OR perforat* OR gangren*) AND

(pediatric* OR child* OR adolescent* OR infant*) AND

(appendectomy OR appendicectomy OR surgery) AND

(antibiotic* OR antimicrobial*) AND

(oral OR "post-discharge" OR "at discharge") AND

(random* OR cohort OR compar* OR trial))

**Scopus =** 93

TITLE-ABS-KEY (appendicitis AND (complicat* OR perforat* OR gangren*) AND

(pediatric* OR child* OR adolescent* OR infant*) AND

(appendectomy OR appendicectomy OR surgery) AND

(antibiotic* OR antimicrobial*) AND

(oral OR "post-discharge" OR "at discharge") AND

(random* OR cohort OR compar* OR trial))

**Ovid =** 12

(exp Appendicitis/ OR appendicitis.tw. OR "complicated appendicitis".tw. OR "perforated appendicitis".tw. OR "gangrenous appendicitis".tw.) AND

(exp Appendectomy/ OR appendectomy.tw. OR appendicectomy.tw. OR "surgical treatment".tw.) AND

(exp Anti-Bacterial Agents/ OR antibiotics.tw. OR "oral antibiotics".tw. OR "post-discharge antibiotics".tw. OR "home antibiotics".tw. OR "discharge prescription".tw.) AND

("no antibiotics".tw. OR "without antibiotics".tw. OR "antibiotic discontinuation".tw. OR "stop antibiotics".tw. OR "antibiotic stewardship".tw.) AND

(exp Pediatrics/ OR pediatric.tw. OR paediatric.tw. OR child.tw. OR children.tw. OR adolescent.tw. OR infant.tw. OR neonate.tw.) AND

(randomized controlled trial.pt. OR cohort.tw. OR "observational study".tw. OR "comparative study".tw. OR "retrospective study".tw. OR "prospective study".tw.)

**Cochrane CENTRAL =** 63

appendicitis AND (complicat* OR perforat* OR gangren*) AND (pediatric* OR child* OR adolescent* OR infant*) AND (appendectomy OR appendicectomy OR surgery) AND (antibiotic* OR antimicrobial*) AND (oral OR "post-discharge" OR "at discharge")
