## Supplementary File 3 for "Post-discharge Oral Home Antibiotics Versus No Oral Home Antibiotics in Complicated Pediatric Appendicitis: A Systematic Review and Meta-analysis"

**Supplementary File 3: Inclusion and Exclusion Criteria**

**Population Inclusion Criteria**

- Pediatric patients (<21 years).
- Diagnosed with complicated acute appendicitis (defined as gangrenous or perforated)
- Managed surgically with appendectomy (either open or laparoscopic)
- Studies reporting outcomes separately for the pediatric population if mixed-age cohorts are included

**Polulation Exclusion Criteria**

- Studies with no surgical intervention (i.e., non-operative management or interval appendectomy).
- Studies that do not include pediatric patients or do not report data separately for this age group.
- Studies conducted exclusively in immunocompromised patients.
- Studies involving patients with abdominal neoplasms or oncologic conditions.
- Studies involving patients with hematologic disorders (e.g., neutropenia, coagulopathies).
- Studies focused exclusively on uncomplicated or chronic appendicitis.

**Studies Inclusion Criteria**

- We will include randomized controlled trials (RCTs), non-randomized controlled trials, and observational cohort studies (both prospective and retrospective) that compare pediatric patients with surgically treated complicated acute appendicitis discharged with versus without oral antibiotic therapy. Only comparative studies with a clearly defined control group (no post-discharge antibiotics) will be included.

**Studies Exclusion Criteria**

- Case reports.
- Duplicate or overlapping studies.
- Retracted studies.
- Reviews, systematic reviews, or clinical guidelines.
- Letters to the editor and editorials.
- Articles published in languages other than English or Spanish.
