## Supplementary material for "Post-discharge Oral Home Antibiotics Versus No Oral Home Antibiotics in Complicated Pediatric Appendicitis: A Systematic Review and Meta-analysis": Table 1

| **Author** | **Country** | **Study design** | **Age** | **Sex M/F** | **Total N** | **Group definitions** | ***N* in IG** | ***N* in CG** | **Discharge criteria and TLC value** | **Antibiotics and posology (type, duration, route)** | **Events by group (IG/CG)** | **p-value** | **Commentaries** |
| --- | --- | --- | --- | --- | --- | --- | --- | --- | --- | --- | --- | --- | --- |
| Desai et al. (2015) [20] | USA | Prospective | **IG**: 9.6(3.8)y^1^  **CG**: 10(3.9)y^1^ | **IG**: 60/210  **CG**: 150/120 | 540 | CAA (PAA): hole in the appendix or fecalith in the abdomen  IG: new protocol  CG: old protocol | **270**  152 discharged before day 5  (135/152 NHA and 17/152 OHA) | **270**  136 discharged before day 5 (136/136 OHA) | **DC:**  -Regular diet tolerance  -Controlled pain with oral medications  -No fever over the previous 12 hours  Preoperative TLC by group:  IG: 17.6 (5.7)^1^  CG: 17.4 (5.8)^1^  (p=0.7)  Predischarge TLC by group:  IG: 8.6 (2.4)^1^  CG: 16.9 (3.5)^1^ (p<0.001) | **Old protocol**:  Preoperative doses of ceftriaxone and metronidazole.  *50 mg/kg (max. 2 gr) IV ceftriaxone and 30 mg/kg (max. 1 gr) IV metronidazole every 24 hours*  -If 5 days IVA: NHA  -If <5 days IVA: 7 days OHA (amoxicillin-clavulanate)  **New** **protocol**:  Preoperative doses of ceftriaxone and metronidazole.  *50 mg/kg (max. 2 gr) IV ceftriaxone and 30 mg/kg (max. 1 gr) IV metronidazole every 24 hours*  -If 5 days IVA: NHA  -If <5 days IVA and leukocytosis: 7 days OHA (amoxicillin-clavulanate)  If <5 days IVA and no leukocytosis: NHA | **IAA**:  IG (global): 12/152  IG (NHA): 11/135  IG (OHA): 1/17  CG (OHA): 6/136 | **IAA (IG vs. CG)**:0.3 | The number of male cases in the study group (60 out of 270) is strikingly low and does not realistically reflect the expected sex-based prevalence of the disease. |
| Anderson et al. (2018) [21] | USA | Retrospective (NSQIP-Pediatric 2015-2016 database) | **IG**: 10.4(4)y^1^  **CG**: 10.1(4)y^1^ | **IG**:1,699/1,099  **CG**: 2,116/1,498 | 6,412 | CAA: visible hole in the appendix, fecalith in the peritoneal cavity outside the appendix, abscess, or diffuse fibrinopurulent exudate in the peritoneal cavity  IG: NHA  CG: IVHA + OHA | **2,798** | **3,614**  OHA: 3,426/3,614  IVHA: 188/3,614 | Preoperative TLC by group:  IG: 16.0 (5.7)^1^  CG:  17.2 (5.3)^1^  (p<0.01)  Predischarge TLC by group: NS | NS | **CPDM:**  IG (NHA): 315/2,798  CG (IVHA+OHA): 522/3,614^a^  **EDV:**  IG (NHA): 269/2,798  CG (IVHA+OHA): 463/3,614^a^  **RA:**  IG (NHA): 153/2,798  CG (IVHA+OHA): 286/3,614^a^  **RO:**  IG (NHA): 17/2,798  CG (IVHA+OHA): 41/3,614^a^  **SSI:**  IG (NHA): 92/2,798  CG (IVHA+OHA): 163/3,614^a^ | **CPDM, EDV, RA:** <0.01  **RO:** 0.03  **SSI:** 0.01 | 188 patients (2.9%) received IVHA instead of OHA, and the analyses did not adequately stratify the OHA group from the IVHA group. The authors were contacted to request the raw data, but no response was received. This should be considered a limitation when interpreting the present study's data.  As this was a retrospective review of a national database, there were no standardized discharge criteria prior to hospital discharge in either group nor was a antibiotics and posology report, which may represent a major source of heterogeneity. |
| Bonasso et al. (2019) [22] | USA | Retrospective | **IG**:9.28y^2^  **CG**:9.37y^2^ | **IG:** 55/42  **CG**:56/26 | 179 | CAA: GA, PAA  IG: NHA CG: OHA | **97**  **NHA**: 69/97  **OHA**: 28/97 | **82**  NHA: 7/82  OHA: 75/82 | **DC:**  -Diet tolerance  -Controlled pain with oral medications  -No fever (<38.5°C)  Preoperative TLC by group: NS  Predischarge TLC by group: NS | **Old protocol**:  Preoperative doses of ceftriaxone and metronidazole.  *50 mg/kg IV ceftriaxone and 30 mg/kg IV metronidazole every 24 hours*  -If <10 days IVA: OHA until 10 days of ATB treatment completion^b^  **New** **protocol**:  Preoperative doses of ceftriaxone and metronidazole.  *50 mg/kg IV ceftriaxone and 30 mg/kg IV metronidazole every 24 hours*  -If TLC was elevated: OHA until 7 days of ATB treatment completion^b^  -If normal TLC: NHA | **IAA:**  CG (global): 8/82  IG (Global): 10/97  OHA: 7/75  NHA: 1/7  **RA**:  CG (global): 11/82  IG (global): 12/97  OHA: 7/28  NHA: 1/69 | **RA**: 1 | Although the authors reported 9 additional intra-abdominal abscesses in the CG group and 9 in the IG group (all corresponding to patients who remained hospitalized and continued on IV antibiotic therapy), these cases were excluded from the group-specific event counts (global), as they are not within the scope of the present review.  The authors report 17 abscesses in the CG and 19 in the IG, with 9 inpatient abscesses in both groups. However, for the IG, the total counts are inconsistent: they report 19 abscesses, of which 9 occurred during hospitalization, 7 in the OHA subgroup, and 1 in the NHA subgroup, resulting in a discrepancy of 2 cases. We contacted the authors to clarify this issue, but did not receive a response.  Although n=28 is inferred for the OHA subgroup (total N=97 minus n=69 discharged without antibiotics, per author's Table 1, the original study's Table 1 uses n=20 for calculating the mean duration of home oral antibiotics, indicating an internal data inconsistency. |
| Rossidis et al. (2020) [23] | USA | Retrospective | **IG**:12(9-14)y^3,c^  **CG**:11(8-14)y^3,c^ | **IG**: 489/325^c^  **CG**: 448/300^c^ | 1,562^c^  (PAA: 338) | PAA: a visible hole in the appendix, a free fecalith, diffuse peritonitis,  or an abscess at the time of surgery  **IG**: new protocol  CG: old protocol | **814**^c^  (PAA: 175)  OHA: 59/175  NHA: 116/175 | **748**^c^  (PAA: 163)  OHA: 145/163  NHA: 18/163 | **DC:**  -Diet tolerance  -Controlled pain with oral medications  -No fever (**≥** 24 hours)  Preoperative TLC by group: NS  Predischarge TLC by group: NS | **Old protocol**:  Postoperative ATB 7-14 days (empirically)  Most patients with OHA (empirically)  **New** **protocol**:  NHA  **IG**: 59/175 OHA  **CG**: 145/163 OHA | **RA:**  CG **(**global): 5/163^d^  IG **(**global): 9/175^d^  **SSI:**  CG **(**global): 11/163^d^  IG **(**global): 8/175^d^  **EDV**:  CG **(**global): 13/163^d^  IG **(**global): 18/175^d^  **RO**:  CG **(**global): 1/163^d^  IG **(**global): 2/175^d^ | **RA:** 0.42  **EDV:** 0.57  **SSI**: 0.48  **RO**:  0.99 | Although it is understood that the study refers to OHA rather than IVHA, this is not explicitly stated at any point in the manuscript. The authors were contacted to confirm this information, but they did not respond.  The authors did not report having determined TLC before discharge.  The article provides overall data for the old protocol and new protocol groups, but does not report raw data for patients with OHA and NHA. We contacted the corresponding author to clarify this information, but did not receive a response. |
| Ferguson et al. (2020) [24] | USA | Retrospective | **IG**:9.7(3.9)y^1^  **CG**:10.1(3.9)y^1,^ | **IG**: 120/93  **CG:**  258/146 | 617 | PAA: intra-  operative visualization of a hole in the appendix, intra-  abdominal stool, or intra-abdominal fecalith  **IG**: old protocol  **CG**: new protocol | **213**  OHA: 12/213  NHA: 201/213 | **404**  OHA: 397/404  NHA: 7/404 | **DC:**  -Diet tolerance  -Controlled pain with oral medications  -No fever (**≥** 24 hours)  -No unexpected abdominal pain  -No emesis  -Normal TLC  Preoperative TLC by group:  IG: 17.9 (5.7)^1^  CG:  17.5(5.3)^1^  (p=0.42)  Predischarge TLC by group: NS | **Preoperative ATB**  *Piperacillin-tazobactam 100 mg/kg/dose IV every 8 h (<40 kg) or 3.375 g IV every 6h (40 kg)*  *Or*  *Cefepime 50 mg/kg/dose IV every 12 h,*  *(max. 2 g/dose)*  *Plus*  *Metronidazole 10 mg/kg/dose IV every 8 h*  *(max. 500 mg/dose)*  **Postoperative ATB**  *Amoxicillin-clavulanate 25-45 mg/kg/d PO divided every 12 h (<40 kg) or 500-875 mg/ dose every 12 h (40 kg)*  **Old protocol (IG)**: no OHA standardization  (12/213 OHA, 201/213 NHA)  **New protocol (CG):** OHA (7 additional days) to all patients at discharge  (397/404 OHA, 7/404 NHA)  *Amoxicilin-clavulanate (n=382), ciprofloxacin + metronidazole (n=13), others (n=2)* | **IAA**  IG (global): 19/213  CG (global): 18/404  **SSI:**  IG (global): 2/213  CG (global): 3/404  **EDV**  IG (global): 10/213  CG (global): 15/404  **RA**^e^  IG (global): 28/213  CG (global): 26/404  **RO**^e^  IG (global): 2/213  CG (global): 2/404 | **IAA**: 0.03  **SSI**: 0.8  **RO**: 0.51  **EDV**: 0.56  **RA**: 0.005 | Patients who underwent conversion to open surgery, as well as those who had primary open appendectomy or remained hospitalized for 8 or more days, were excluded, which may introduce a potential selection bias. For the purposes of this review, the pre-standardization group has been defined as the intervention group (IG) and the post-standardization group as the control group (CG), since the authors standardized the practice of prescribing, rather than withholding, OHA.  The article provides overall data for the old protocol and new protocol groups, but does not report raw data for patients with OHA and NHA. We contacted the corresponding author to clarify this information, but did not receive a response.  Patients were excluded if an open or laparoscopic-converted-to-open appendectomy was performed. |
| Gordon et al. (2020) [25] | USA | Retrospective | **IG:** 8.0 (6.0-11)y ^3^  **CG:** 9.0 (6.0-12)y ^3^ | 157/96 | 253 | CAA (PAA): Perforation of the appendix without abscess or phlegmon before surgery or identification of a perforated appendix intra- operatively  IG: new protocol (NHA)  CG: old protocol (OHA) | 91 | 162 | **DC:**  -Afebrile for at least 24 h  -Tolerating a regular diet  -Minimal abdominal tenderness  -Normal TLC  Preoperative TLC by group:  IG: 16.6(13.9-19.7)^3^  CG:  16.8(13.8-20.3)^3^  (p=0.57)  Predischarge TLC by group:  IG: 8.1(6.8-10.4)^3^  CG:  9.5(7.4-10.9)^3^  (p=0.02) | **Old protocol:**  While admitted: IV piperacillin/tazobactam  or IV meropenem  OHA (ciprofloxacin/metronidazole or amoxicillin/clavulanate)  **New protocol:**  While admitted: IV piperacillin/tazobactam or IV meropenem  NHA | **IAA**  IG (NHA): 7/91  CG (OHA): 14/162  **RA**  IG (NHA): 10/91  CG (OHA): 17/162 | **IAA:** 0.99  **RA:** 0.99 | Although all patients who underwent appendectomy before November 2017 were discharged on OHA, the length of the antibiotic course was not standardized. This varied from 5 to 14 d based on the discretion of each surgeon.  In addition, patients who were sent home without antibiotics (NHA) had significantly lower TLC counts at the time of discharge (p=0.02). |
| Ketha et al. (2021) [26] | USA | Retrospective | **IG**: 8.8y^2^  **CG:** 9y^2^ | 72/57 | 129 | PAA: clinically perforated appendicitis at time of surgery as noted on operative reports.  IG: new protocol (NHA)  CG: old protocol | 58  (OHA:0/58  NHA: 58/58) | 71  (OHA: 16/71  NHA: 55) | DC:  -Afebrile for 24 h (<38.5 C)  - Adequate diet tolerance.  - Adequate pain control with oral medications.  Preoperative TLC by group: NS  Predischarge TLC by group: NS | **Old protocol:**  Preoperative: ceftriaxone 50 mg/kg and metronidazole 30 mg/kg IV  While admitted: ceftriaxone 50 mg/kg and metronidazole 30 mg/kg IV once daily  If DC accomplished before day 7, TLC was checked. If TLC >13.5x10^9^/L, OHA (amoxicillin/clavulanic acid) 7 days  If TLC ≤ 13.5x10^9^/L, NHA  **New protocol:**  Preoperative: ceftriaxone 50 mg/kg and metronidazole 30 mg/kg IV  While admitted: ceftriaxone 50 mg/kg and metronidazole 30 mg/kg IV once daily  NHA | **IAA**  IG (NHA): 7/58  CG (global): 9/71  **RA**  IG (NHA): 10/58  CG (global): 9/71 | **IAA:** 1  **RA:** 0.61 | Doesn´t specify if the abscess occurs post-discharge  The article provides overall data for the old protocol and new protocol groups but does not report raw data for patients with OHA and NHA. We contacted the corresponding author to clarify this information but did not receive a response. |
| Plattner el al. (2021) [27] | USA | Retrospective | 10.3 (3.9) y^1^ | 56%/44%  186/147^g^ | 333  (295)^f^ | **PAA:** Presence of intra-luminal contents outside of the bowel at time of surgery (as per the operative note)  IG: NHA  CG: OHA, IVHA, OHA+IVHA | NHA:115 | 180  (OHA, IVHA, OHA + IVHA)  OHA: 135/180 | **DC:** NS  **TLC:** NS  Preoperative TLC by group: NS  Predischarge TLC by group:  IG: 9^2^  CG: 10.6^2^  (p<0.001) | **Broad-spectrum antibiotic coverage**:  meropenem, piperacillin-tazobactam, and/or a fourth-generation cephalosporin.  **Narrow-spectrum antibiotic coverage**:  metronidazole plus a third-generation cephalosporin, most commonly ceftriaxone  **IG**: NHA  **CG**: OHA, IVHA, OHA+IVHA | **IAA**  IG (NHA): 14/115^g^  CG (OHA+IVHA+OHA/IVHA): 38/180^g^  CG (OHA): 12/135^g^  **RA**  IG (NHA): 9/115^g^  CG (OHA+IVHA+OHA/IVHA): 14/180^g^  CG (OHA): 11/135^g^ | **IAA:**  OHA+IVHA+OHA/IVHA: <0.01  OHA vs. NHA:  0.79  **RA:**  OHA vs. NHA:  0.39 | The study included 333 patients, but only 295 underwent appendectomy.  The choice between narrow and broad-spectrum antibiotic coverage was at surgeon´s discretion  Initial broad-spectrum antibiotic use was associated with increased odds of postoperative abscess formation (OR 3.17, 95%CI 1.58-6.38)  Patients with home antibiotics (OHA, IVHA and mixed OHA + IVHA) had a significantly higher rate of postoperative abscess formation than NHA while patients with OHA did not differ in abscess formation compared to the NHA group.  The average discharge TLC was significantly higher in the group of patients who experienced readmission compared to those who did not (p<0.001). |
| Anandalwar  et al. (2021) [28] | USA | Retrospective (NSQIP-P database from January 2013 to June 2015) | 10 (7-17)  y^3^ | Before PM:  416/370  After PM:  334/240 | Before PM: 711 (786)  PM: 574 | **CAA:** Presence of a visible hole, diffuse fibrinopurulent exudate extending outside the right lower quadrant and pelvis (defined as exudate in more than two quadrants of the abdomen and pelvis), intra-abdominal abscess, or an extra-luminal fecalith    **IG:** NHA  **CG:** OHA | Before PM:  306  After PM: 287 | Before PM:  405  After PM: 287 | **DC:** NS  Preoperative TLC by group (before PM):  IG: 74 (24.2)^5^  CG: 91 (22.5)^5^  (p=0.02)  Preoperative TLC by group (after PM):  IG: 72 (25.1)^5^  CG: 77 (26.8)^5^  (p=0.89)  Predischarge TLC by group: NS | NS | **OSI (before PM)^h^:**  IG (NHA):  6.2%  19/306^g^  CG (OHA):  4.4%  18/405^g^  **OSI (after PM)^h^:**  IC (NHA):  6.6%  19/287^g^  CG (OHA):  4.2%  12/287^g^  **RA (after PM):**  IC (NHA):  5.9%  17/287^g^  CG (OHA):  5.6%  16/287^g^ | **OSI (before PM)**: 0.30  **OSI (after PM:** 0.21  **RA (after PM):** 0.71  **IAA:** 0.30 | Since this work reports data from a database that includes 29 NSQIP-Pediatric hospitals, therefore it can´t be assumed that discharge criterion, and the antibiotic used and its posology was consistent among all the hospitals, as it has not been specified on the paper.  Adjusted analyses (after PM) showed no statistically significant differences between groups (OHA/NHA) for either OSI or RA.  The sex distribution table reflects the pre-exclusion cohort (n=786), which explains why male + female totals exceed the final sample. After excluding patients with inpatient imaging or IV antibiotics at discharge, the analytic cohort was reduced to 711 |
| Jen et al. (2022) [29] | USA | Retrospective | Main study arm (no initial DSI):  IG: 9.9 (4.0)y^1^  CG: 10.2 (4.2)  y^1^ | 232/173  Main study arm (no initial DSI):  203/160 | 405  Main study arm (no initial DSI):363 | **CAA:** (PAA): either intraoperative or pathologic findings of PAA  **IG**: NHA  **CG**: OHA | 288  Main study arm (no initial DSI):  277 | 117  Main study arm (no initial DSI):  86 | **DC:** based on attending surgeon preference but based on somewhat standardized guidelines that included remaining afebrile for at least 24 h, tolerating a regular diet, and having pain that was well-controlled with oral analgesics.  Preoperative TLC by group:  IG: 16.9(5.3)^1^  CG:  17.5(6.4)^1^  (p=0.36)  Predischarge TLC by group: NS | **Initial antibiotics**  Ceftriaxone/metronidazole (CG,n=75; IG, n=259)  Piperacillin/tazobactam (CG, n=6; IG, n=8)  Other (CG, n=5; IG, n=10) **OHA (CG):**  Ciprofloxacin/metronidazole (n=72)  Amoxicillin-clavulanate (n=12)  Other (n=2) | **OSI:**  IG (NHA): 9/277  CG (OHA): 4/86  **EDV:**  IG (NHA): 23/277  CG (OHA): 10/86  **RA:**  IG (NHA): 10/277  CG (OHA): 6/86  **SSI:**  IG (NHA): 5/277  CG (OHA): 0/86 | **OSI:**  0.54  **EDV:**  0.35  **RA:** 0.18  **SSI:**  0.21 | The authors do not analyze the presence of OSI, EDV, RA or SSI based on the antibiotics administered. The use of multiple different antibiotics and regimens without protocol may constitute a relevant source of heterogeneity. |
| Morita et al. (2022) [30] | Japan | Retrospective (Diagnosis Procedure Combination database) | 3-18y^4^ | Before PM:  7,842/5,258  After PM:  3,330/2,208 | Before PM:  13,100  After PM:  5,538 | **CAA:** NS  IG: NHA  CG: OHA | Before PM:  9,599  After PM:2,769 | Before PM:  3,501  After PM:2,769 | **DC:** NS  Preoperative TLC by group: NS  Predischarge TLC by group: NS | NS | **RA due to OSI (before PM):**  IG (NHA): 333/9,599  CG (OHA): 163/3,501  **RA due to OSI (after PM):**  IG (NHA): 93/2,769  CG (OHA): 145/2,769  **Global RA (within 60 days of discharge) (before PM):**  IG (NHA): 549 /9,599  CG (OHA): 247/ 3,501  **Global RA (within 60 days of discharge) (after PM):**  IG (NHA): 153/2,769  CG (OHA): 205/2,769 | **RA due to OSI (before PM):** 0.002  **RA due to OSI (after PM**: 0.001  **Global RA (before PM):** 0.005  **Global RA (after PM):** 0.004 | Since this work reports data from a nationwide database that includes more than 1,200 hospitals, therefore it can´t be assumed that discharge criterion, and the antibiotic used and its posology was consistent among all the hospitals, as it has not been specified on the paper. |
| Theodorou et al. (2022) [31] | USA | Retrospective | **Old protocol**: 8.9 (6.2-11.4) y^3^  **New protocol**: 9.2 (6.7-12.3)  y^3^ | 132/78 | 210 | **CAA:** PAA: hole in the appendix, a fecalith in the abdomen, with or without an associated abscess or purulent fluid.  **IG**: New protocol  **CG**: Old protocol | 97  OHA:  14/97 | 113  OHA: 80/113 | **DC:**  -Afebrile for 24 h  - Tolerating a diet  -Pain manageable with oral medications  -A benign examination  Preoperative TLC by group:  IG: 16.3(14-18.2^3^  CG:  18.5(14.8-21.9^3^  (p=0.003)  Predischarge TLC by group:  IG: 9.2(7.3-10.7)^3^  CG:  8.9(7.7-10.9)^3^  (p=0.77) | Preoperative administration of intravenous IV ceftriaxone and metronidazole  **Postoperative treatment:** NS  **Old protocol:**  If normal TLC: 5 days OHA  If abnormal TLC: 10 days OHA  OHA: 80/113; NHA: 33/113  **New protocol**  If normal TLC: NHA  If abnormal TLC: 10 days OHA  OHA: 14/97; NHA: 83/97 | **SSI:**  IG (global): 9/97  CG (global): 2/113  NHA: 8/116^g^  OHA: 3/94^g^  **RA:**  IG (global): 11/97  CG (global): 6/113 | **SSI (CG vs. IG):**  0.03  **SSI (OHA vs. NHA)**: 0.53  **RA**:  0.13 | Open appendectomy patients were excluded as there may be a higher rate of surgical site infections in this patient population compared to laparoscopic appendectomy  The authors do not specify a cut-off point for TLC.  There were no significant differences in TLC at discharge between the SSI and non-SSI groups.  The article provides overall data for the old protocol and new protocol groups, but does not report raw data for patients with OHA and NHA. We contacted the corresponding author to clarify this information, but did not receive a response.  The authors report proportions conditioned on outcome: 3/11 patients with SSI (27.3%) received antibiotics, and 91/199 without SSI (45.7%) did. However, restructuring the data by treatment group shows that 3 of 94 patients who received antibiotics developed SSI (3.2%), compared to 8 of 116 without antibiotics (6.9%). This corresponds to infected:non-infected ratios of 0.03 (3/91) in the antibiotic group and 0.07 (8/108) in the no-antibiotic group—identical to the original data but correctly aligned for effect estimation. |
| Russell et al. (2023) [32] | USA | Retrospective (NSQIP database from January 2015 to May 2022) | **Old protocol**:  9.8(4)  y^1^  **New protocol**:  9.6(4)  y^1^ | 173/133^g^ | 306 | **CAA:** Presence of a visible hole, diffuse fibrinopurulent exudate (defined as exudate in >2 quadrants of the abdomen and pelvis), intra-abdominal abscess, or an extraluminal fecalith  **IG**: new protocol  **CG**: old protocol | 121  OHA:10/121 NHA:111/121 | 185  OHA:170/185^g^  IVHA:1/185^g^ NHA:14/185^g^ | **DC:**  -Adequate oral intake  -Feeling generally well.  -No fever (<38.5ºC)  Preoperative TLC by group:  IG: 17(5.9)^1^  CG:  17.7(6.2)^1^  (p=0.24)  Predischarge TLC by group: NS | In both protocols, patients were treated with intravenous (IV) Ceftriaxone and IV Metronidazole while hospitalized.  CG: The CG was discharged home on OHA (Augmentin or Cefdinir and Metronidazole if they had an allergy to Augmentin) if their TLC was elevated at the time of discharge  IG: NHA. No predischarge TLC check. | **SSI:**  IG (global): 3/121  CG (global): 3/185  **OSI**:  IG (global): 9/121  CG (global): 13/185  **RA**:  IG (global): 9/121  CG (global): 11/185 | **SSI:** 0.68  **OSI**: 1  **RA**: 0.64 | The article provides overall data for the old protocol and new protocol groups but does not report raw data for patients with OHA and NHA. We contacted the corresponding author to clarify this information but did not receive a response. |
| Patwardhan  Et al. (2025) [33] | USA | Retrospective (NSQIP-P database from 2021 to 2023) | No-OHA: 9.3 (4.3) y^1^  OHA if high TLC:  10.1 (4.0) y^1^  OHA to complete a minimum number of total postoperative ATB days:  9.5 (4.0) y^1^  Standardized OHA discharge:  10.1 (3.7) y^1^ | 827/515 | 1,342 | **CAA:** Intraoperative findings of CAA: the presence of a visible hole in the appendix, an intraperitoneal fecalith, diffuse fibrinopurulent exudate, or the presence of an abscess.  **IG**: NHA cohort (cohort 1)  **CG**: OHA cohort (cohort 4) | 158  OHA: 7/158  NHA: 151/158 | 64  OHA: 59/64  NHA: 5/64 | **DC:**  **-**Lack of fever  -Resolution or significantly improving abdominal pain  -Tolerance of adequate oral intake  Preoperative TLC by group:  IG: 17.1(6.0)^1^  CG:  17.3(6.2)^1^  (p=0.8)(4-groups comparison)  Predischarge TLC by group: NS | Postoperative antibiotic protocol used at each site was collected, with 6/9 centers using ceftriaxone/metronidazole as  intravenous therapy while inpatient and 7/8 centers using amoxicillin/clavulanic acid as the oral antibiotic of choice on discharge (Original articles` Supplemental Table 1)  **IG**: 7/158 OHA; 151/158 NHA  **CG**: 59/64 OHA; 5/64 NHA | **SSI:**  IG (Global): 9/158  CG (Global): 6/64  **RA**:  IG (Global): 12/158  CG (Global): 3/64 | **SSI (4-protocols comparison):** 0.43  **RA (4-protocols comparison):** 0.06 | Since this work reports data from a database that includes 29 NSQIP-Pediatric hospitals, therefore it can´t be assumed that discharge criterion, and the antibiotic used and its posology was consistent among all the hospitals.  Hospitals were classified into four groups based on their protocol: 1) No OHA, 2) OHA if high TLC on the day of discharge, 3) OHA to complete a minimum number of total post-operative ATB days, and 4) standardized OHA discharge regardless of inpatient antibiotic duration.  Only patients undergoing laparoscopic appendectomy were included.  The article provides overall data for different protocols and new protocol groups but does not report raw data for patients with OHA and NHA. We contacted the corresponding author to clarify this information, but did not receive a response. |

**Table 1. Summary of publications included in this review.**

**NS**: Not specified; **y**: Years.

**DC:** Discharge criteria;	**IG**: Intervention group (no oral antibiotic group); **CG**: Control group (post-discharge oral antibiotic group); **CAA:** Complicated acute appendicitis; **PAA**: Perforated acute appendicitis; **GA**: Gangrenous appendicitis

**IV**: intravenous; **OHA**: oral antibiotics (home); **IVA**: intravenous antibiotics (hospitalization); **IVHA**: intravenous antibiotics (home); **NHA**: no-home antibiotics; **ATB**: Antibiotics;

**IAA**: Intraabdominal abscess (post-discharge); **SSI**: surgical site infection (post-discharge); **CPDM**: Composite post-discharge morbidity; **EDV**: Emergency Department visit; **RA**: Readmission; **RO**: Reoperation; **OSI**: organ-space infection (post-discharge), **DSI**: Deep space infection

**TLC**: Total leukocyte count (WBC)

**gr**: grams

**NSQIP**: National Surgical Quality Improvement Program; **PM**: Propensity matching

**1:** Mean (standard deviation); **2:** Mean; **3**: Median (interquartile range); **4:** range; **5**: Highest quartile, n, (%)

**a**: The CG group includes, without stratification, both patients with oral home antibiotics (n=3,426) and patients with intravenous home antibiotics (n=188); **b**: Although Bonasso et al. use the expression 'sent home with oral antibiotics to complete X days of treatment', which may create ambiguity as to whether 'X days' refers to the total duration of antibiotic therapy including inpatient administration or only the outpatient course, Table 1 of the paper suggests that 'X days' refers to the total duration of treatment; **c**: Rossidis et al. report age and sex distribution data for the overall cohort, without specifying the sociodemographic characteristics of the PAA group, which is the focus of the present study; **d**: The reported data refer to complications by overall group; however, specific complication rates for the OHA group and the NHA group were not provided.; **e**: In the study by Ferguson et al., statistically significant differences in readmission (RA) rates are reported between the IG and CG. However, this includes a broad range of signs and symptoms, and there is considerable variability in the distribution of etiologies between groups. For instance, in the group where OHA administration at discharge was standardized, a higher number of patients were readmitted with nonspecific diagnoses such as 'abdominal pain'. The rate of intra-abdominal abscess (IAA) as the reason for RA was 68% and 69% in the IG and CG, respectively; **f:** In the study by Plattner et al. the total number of patients included was 333 of which only 295 were initially managed surgically; **g:** Data estimated through inferential analysis based on the results reported in the article; **h**: OSI as defined by NSQIP: must meet all three of the following criteria: 1. Infection involves any part of the anatomy (organ/space) that was: Opened or manipulated during the operation, and is not the incision itself (i.e., not superficial or deep incisional). 2 The patient has at least one of the following signs or symptoms: Fever (>38°C), localized pain or tenderness and other clinical evidence of infection. 3. One of the following must be true: Purulent drainage from a drain placed into the organ/space, organism isolated from a culture of fluid/tissue from the organ/space, abscess or other infection found during reoperation, direct exam, histopathology, or imaging, or diagnosis of an organ/space infection made by a physician;
